# Functional dynamic network connectivity differentiates biological patterns in the Alzheimer’s disease continuum

**DOI:** 10.1101/2024.10.14.24314934

**Authors:** Lorenzo Pini, Lorenza Brusini, Alessandra Griffa, Federica Cruciani, Gilles Allali, Giovanni B Frisoni, Maurizio Corbetta, Gloria Menegaz, Ilaria Boscolo Galazzo, the Alzheimer’s Disease Neuroimaging Initiative

**Affiliations:** Padova Neuroscience Center, University of Padova, Via Giuseppe Orus 2/B, 35131 Padova, Italy; Department of Neuroscience, University of Padova, via Belzoni 160, 35121 Padova, Italy; Department of Engineering for Innovation Medicine, University of Verona, Strada le Grazie 15, 37134 Verona, Italy; Leenaards Memory Center, Department of Clinical Neurosciences, Lausanne University Hospital and University of Lausanne, Montpaisible 16, 1011 Lausanne, Switzerland; Medical Image Processing Laboratory, Neuro-X Institute, École Polytechnique Fédérale De Lausanne (EPFL), Campus Biotech Chemin des Mines 9, 1202 Geneva, Switzerland; Memory Clinic and LANVIE - Laboratory of Neuroimaging of Aging, University Hospitals and University of Geneva, Rue du Général-Dufour 24, 1211 Genève, Switzerland; Veneto Institute of Molecular Medicine, VIMM, Via Giuseppe Orus 2, 35129 Padova, Italy

**Keywords:** Dynamic connectivity, molecular pathology, brain networks

## Abstract

Alzheimer’s disease (AD) can be conceptualized as a network-based syndrome. Network alterations are linked to the molecular hallmarks of AD, involving amyloid-beta and tau accumulation, and neurodegeneration. By combining molecular and resting-state functional magnetic resonance imaging, we assessed whether different biological patterns of AD identified through a data-driven approach matched specific abnormalities in brain dynamic connectivity. We identified three main patient clusters. The first group displayed mild pathological alterations. The second cluster exhibited typical behavioral impairment alongside AD pathology. The third cluster demonstrated similar behavioral impairment but with a divergent tau (low) and neurodegeneration (high) profile. Univariate and multivariate analyses revealed two connectivity patterns encompassing the default mode network and the occipito-temporal cortex, linked respectively with typical and atypical patterns. These results support the key association between macro-scale and molecular alterations. Dynamic connectivity markers can assist in identifying patients with AD-like clinical profiles but with different underlying pathologies.

Within the clinical continuum of Alzheimer’s disease (AD), we identified two main groups: one characterized by typical AD neuropathological changes, and the other by atypical pathophysiological mechanisms. Univariate and multivariate analyses revealed two dynamic functional connectivity patterns, involving the default mode network and the occipito-temporal cortex, respectively.

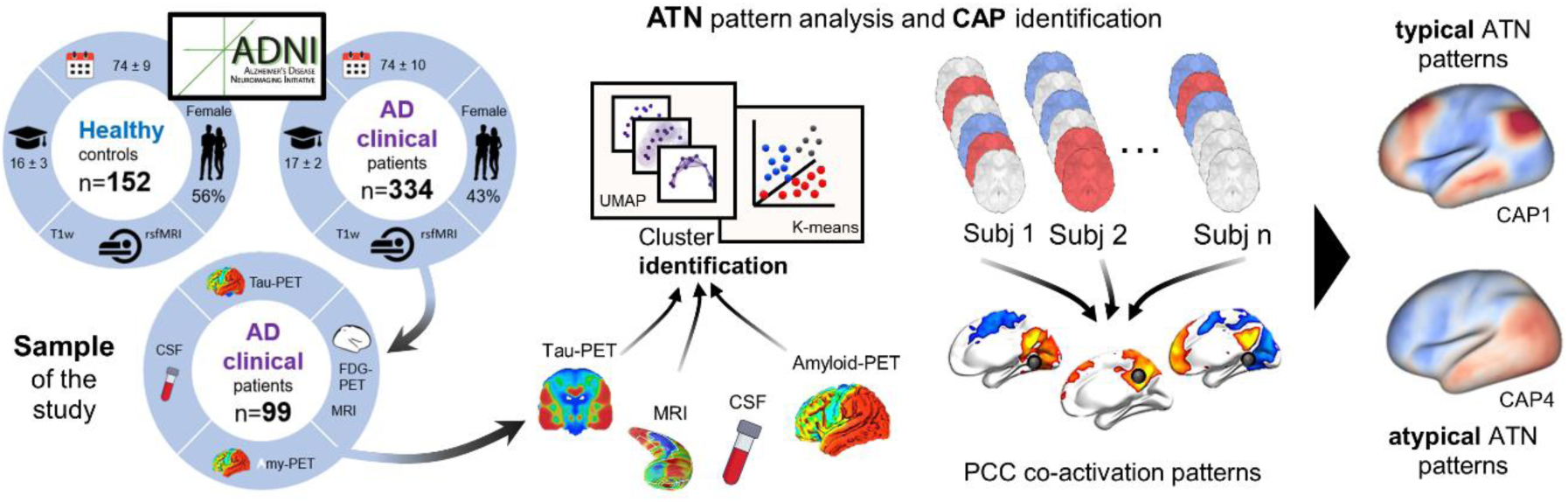

## INTRODUCTION

An operational framework, known as ATN from the biological markers of (A)myloid-beta (Aβ), (T)au, and (N)eurodegeneration, has facilitated a transition from a clinical perspective to a biological viewpoint of Alzheimer’s disease (AD), decoupling diagnosis from discrete clinical stages.^1^ Classification into ATN stages requires the designation of individuals as positive (+) or negative (-) for Aβ, tau, and neurodegeneration (A+/A−, T+/T−, N+/N−). While Aβ thresholds for positivity are relatively consistent across amyloid positron emission tomography (PET) studies, there is methodological variance in determining tau cut-offs based on PET, leading to limited consensus.^2^ In a recent review, Weigand et al. (2022) found that tau PET quantifications were consistently reliable across studies, but dichotomization cut-points varied significantly. This variability, with cut-points ranging from 1.13 to 2.79, may be due to differences in sample characteristics and processing methodologies.^2^ Similarly, there is no universal cut-off value to define abnormal Aβ cerebrospinal fluid (CSF), partially due to the variability of measurements across laboratories.^3^

Overall, performance findings in thresholding methods, either based on CSF or PET data, show some variability mainly due to differences in methodology, such as processing steps, region-of-interest selection and different statistical approaches.^2,4^ Additionally, ATN cut-off points are not sensible to individual variability (e.g., age, genetics, sociodemographic) or specific brain regions, making challenging to establish universal sensitive/specific values.^5–7^ These limitations appear more pronounced in view of non-AD pathophysiological mechanisms involving ATN markers, such as limbic-predominant age-related TAR DNA binding protein 43 (TDP-43) encephalopathy (LATE)^8^, mimicking both AD clinical and N phenotypes.^9,10^

Along with such complexity, the ATN poses also challenges for individuals with a clinical AD diagnosis but atypical biomarker abnormalities, such as amyloid negative patients with positive marker of tau and/or degeneration (A−T+N−, A−T−N+ and A−T+N+), all profiles referring to the suspected-non-AD pathology (SNAP). The underlying pathology of SNAP can be highly heterogeneous, although the prevalence of these possible combinations is still an open question, underling the necessity of new insights into the pathophysiology of SNAP.^11^

Taking all these aspects together, given the challenges posed by the definition of universal cut-offs, inter-individual variability, overlapping pathophysiological mechanisms (e.g., LATE) and unclear ATN profiles (e.g., SNAP), data driven approaches rather than strict cut-off points may aid in differentiating the biological patterns in patients with a clinical diagnosis of AD. The core ATN system could also be extended by adding further brain fingerprints serving as novel biomarkers, such as functional connectivity (FC) from resting state functional magnetic resonance imaging (rs-fMRI). Such technique measures Brain signal arising from neural activity-related local blood flow changes and the subsequent alteration of the oxy-to deoxyhemoglobin ratio, allowing to detect large-scale neural networks. These circuits exhibit specific spatiotemporal features and a certain degree of correspondence with behavior.^12^ Among the well-known resting-state networks, the default mode network (DMN) has been shown to be highly vulnerable in AD,^13,14^ to be linked with memory deficits,^13,15^ and to predict brain Aβ and tau deposition, suggesting that the molecular pathology may spread along functional pathways^16,17^. However, it is unlikely that there is a 1:1 ratio between pathology and distinctive network alterations. Imbalances within and between networks could better predict clinical and cognitive impairment, rather than focusing solely on a single network connectivity breakdown.^18^ Moreover, exploring dynamic FC (dFC) changes rather than static brain connectivity can provide a finer-grained characterization of FC modulations. Among these approaches,^19^ methods based on the frame-wise description of rs-fMRI time courses, e.g., co-activation patterns (CAPs) analysis, are gaining increasing interest thanks to their ability to generate voxel-wise brain states representing how a specific brain region recurrently co-activates or co-deactivates with all the others. Previous studies applied CAPs in both healthy and psychiatric disorders,^20,21–23,24^, showing higher sensitivity to characterize connectivity reconfiguration.

Based on these assumptions, we first aimed at identifying individuals sharing similar biological ATN patterns in the clinical continuum of AD, by relying on a data-driven machine learning based approach. We hypothesize this would allow to detect groups exhibiting diverse ATN patterns which may correspond to different temporal stages of typical AD as well as atypical patterns suggestive of distinct pathological mechanisms.

We then aimed at characterizing the dFC and associated brain states to capture the time-varying brain activity and disease-induced modulations. We hypothesized that different dFC patterns would emerge in the ATN-based subgroups, providing a complete characterization of the identified ATN profiles, from a biological to physiological perspective. Moreover, we evaluated whether the dFC temporal measures associated with the different brain CAPs could hold predictive power in discriminating patients with diverse biological hallmarks. All these elements can uncover the association between data-driven ATN profiles and specific dFC fingerprints, reinforcing the assumption that manifold ATN profiles align with dFC.

## METHODS

### Participants and dataset

From the Alzheimer’s Disease Neuroimaging Initiative (ADNI), we included 334 MCI/AD patients (74±10 years, female 43%) and 152 cognitively unimpaired individuals (CU)(74±9 years, female 56%). For up-to-date information, see www.adni-info.org. The study was carried out in accordance with the guidelines of the Declaration of Helsinki.

Data selection was based on the availability of structural T1-weighted (sMRI) and rs-fMRI images acquired on 3T scanners (**Table 1**). This cohort (referred to as the “imaging cohort”) was used to compute CAPs. A subset of n=99 patients with fully availability of specific ATN measures (mean age 74 ± 8, female 51%; “ATN cohort”) was considered for further analyses. Specifically, ADNI individuals with the availability of the following data were considered as the “ATN cohort”: 1) Aβ_42_, total tau and phosphorylated tau (ptau) levels assessed in the CSF; 2) regional caudal middle frontal (CMF) gyrus, hippocampus, entorhinal cortex (EC), and amygdala mean values from both tau-PET and amyloid-PET standardized uptake values (SUVR); 3) whole brain composite tau-PET and amyloid-PET values, along with normalized FDG-PET signal within a set of predefined and previously validated regions of interest (metaROIs).^25^ Only participants with these data fully available and collected within 1 year from the MRI exam were included in the ATN cohort, to avoid any potential biases deriving from missing data. Finally, regional grey matter volumes of the hippocampus, amygdala, EC, and CMF gyrus were computed from the minimally preprocessed bias-field corrected sMRI data (fsl_anat, fsl.fmrib.ox.ac.uk/fsl/fslwiki) based on the Desikan-Killiany atlas and using FreeSurfer v.6 (surfer.nmr.mgh.harvard.edu/). These regions were select to cover regions showing alterations in both AD neuropathological change (AD-NC) and not AD-NC mimicking AD (e.g., LATE)^26^.

**Table 1.**
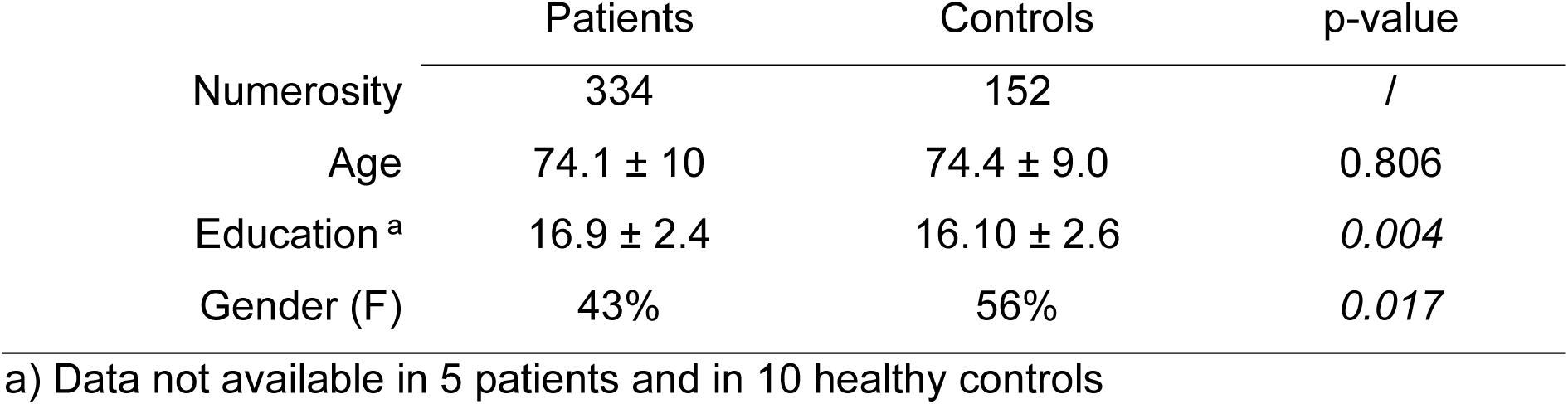
Main sociodemographic characteristics of patients and controls included in the study.

|  | Patients | Controls | p-value |
| --- | --- | --- | --- |
| Numerosity | 334 | 152 | / |
| Age | 74.1 ± 10 | 74.4 ± 9.0 | 0.806 |
| Education <sup>a</sup> | 16.9 ± 2.4 | 16.10 ± 2.6 | *0.004* |
| Gender (F) | 43% | 56% | *0.017* |

a) Data not available in 5 patients and in 10 healthy controls

Volumetric features were normalized to the respective total intracranial brain volume and averaged across the hemispheres. ATN measures were z-scored with respect to the mean and standard deviation values estimated from the whole group of 152 CU to depict variations compared to normative data. CSF Aβ_42_ values were multiplied by -1 to align with amyloid PET measures as elevated CSF Aβ_42_ indicates normal patterns, while high amyloid PET values signify severe amyloid pathology. Besides age, education level and gender, for each patient of the ATN cohort cognitive and clinical information were retrieved: Mini-Mental State Examination (MMSE) score, Clinical Dementia Score – Sum of Boxes (CDR-SB), total score from Alzheimer’s Disease Assessment Scale-13 (ADAS-13), and composite measures computed from the ADNI consortium for memory, executive functions, language, and visuospatial abilities.^27,28^

### Co-activation patterns analysis

Rs-fMRI data from the *imaging cohort* were preprocessed using the FMRIB Software Library (version 6.0). Details are reported in the **Supplementary Material** (paragraph S1.1). The denoised data, all truncated at the 192^nd^ volume to ensure the same amount of data was available in all subjects, were then used for the CAPs analysis, conducted using the TbCAPs toolbox^29^ on MATLAB v2021a. The toolbox was adapted with in-house scripts to deal with large amount of data. The following steps were performed, starting from CU: 1) Z-score transformation of the rs-fMRI signals; 2) data masking using TbCAPs default grey matter mask; 3) frames with framewise displacement (FD)>0.1 were excluded to control for motion; 4) a single seed (PCC) was selected using a spherical region of interest (ROI) of 5-mm radius centered in the MNI peaks previously described (x=0, y=56, z=16).^30^ PCC was chosen according to its key role as hub of the functional connectome and due to its centrality within the memory network systems vulnerable in neurodegenerative disorders;^31^ 5) volumes with PCC signal intensity in the top 30% were retained (positive only) and considered as PCC-activated frames. Spatial clustering was performed on the extracted PCC frames to compute CAPs. The selection of the optimal number of clusters was determined using the consensus clustering algorithm (2): k-means clustering was run across 20 folds for cluster numbers (K) ranging from 3 to 6, randomly selecting 80% of the data for each fold. The K-range was selected according to a previous study investigating PCC-CAPs and reporting K=5 as the optimal value (3). The first four steps were performed also in patients’ data. However, their CAPs were not directly computed but each PCC-activated frame was assigned to one of the CAPs estimated in the CU control group, based on the distribution of the spatial correlation values between the CU PCC frames and the considered CAP. If the spatial correlation value of the patient frame with the given CAP exceeded the 5th percentile of this distribution, the frame-to-CAP assignment was performed, otherwise it was left unassigned. To check whether this assignment generated comparable CAPs between CU and patients, a spatial correlation between the group-averaged CAP was performed.

For each CAP and patient, the following temporal metrics were computed: i) resilience; ii) betweenness centrality; iii) average duration; iv) subject entries; v) counts; vi) out degree; vii) transition probabilities. Finally, we computed for each subject a frame-wise CAPs average. These spatial maps correspond to the spatial distribution of the fMRI signal grouped into specific CAPs. A full description of CAPs metrics is reported in the **Supplementary Material** (paragraph S1.2). A general overview of the methodology is reported in **Fig. 1**.

**Figure 1.**
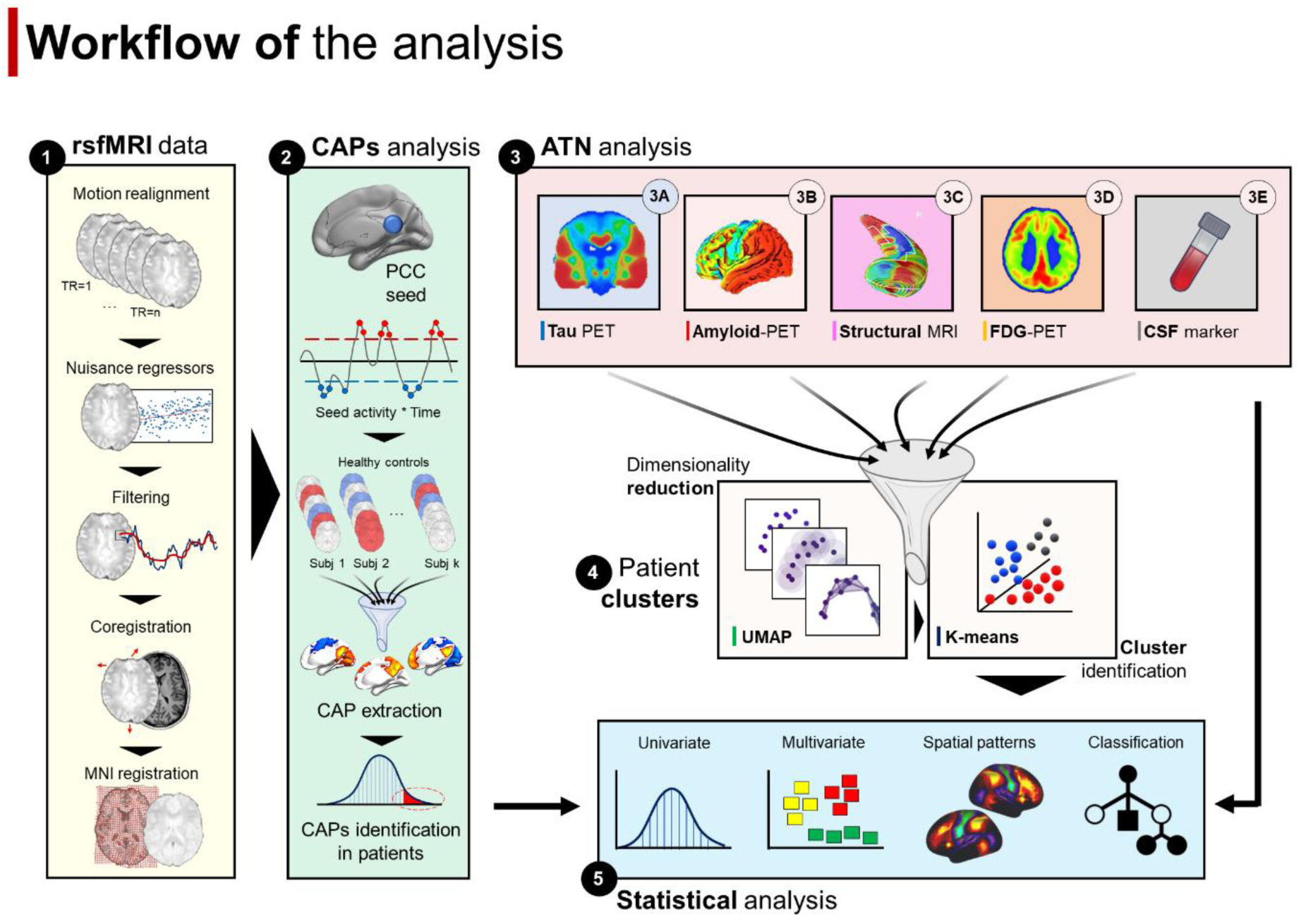
Workflow of the analysis. Panel 1: Resting-state fMRI data (rsfMRI) were preprocessed and (panel 2) co-activation patterns (CAPS) extracted from the whole cohort of controls and then projected to the patient’s cohort. Panel 3: molecular and anatomical data from patients were selected, including tau-PET outcomes (3A), amyloid-PET data (3B), volume measures (3C), brain metabolism (3D), and CSF values for amyloid, total tau and ptau (3E). Panel 4: dimensionality reduction (UMAP) and K-means clustering were performed in patients, identifying three main sub-groups within the clinical AD continuum. Panel 5: A complete set of statistical analyses were performed to assess differences in both ATN and CAPs metrics across patients’ clusters.

### ATN data-driven analysis and patient clustering

For the subgroup of patients included in the *ATN cohort* a data-driven analysis was performed on the ATN measures. Before clustering, the z-scored ATN measures were processed with the Uniform Manifold Approximation and Projection (UMAP), a non-linear embedding approach that distributes data variability along major axes.^32^ We employed UMAP for its ability to transform data onto a newly constructed manifold while maintaining the original pairwise distances between data points on a global scale. Thus, patients with similar ATN profile cluster together in the UMAP space, while patients with different distribution of ATN measures are located further apart. Further, we performed a sensitivity analysis to ensure the robustness of these clusters (see paragraph S1.3 in the **Supplementary Material** for further details). Overall, this step allowed to identify specific clusters based on the ATN measures.

### Statistical analysis

Statistical analyses were run to investigate differences in the ATN, clinical, cognitive, and CAPs’ temporal features between patient clusters identified through the UMAP-Kmeans algorithm, using Python 3.9.

#### Comparing biological, clinical, and cognitive measures between patient clusters

For ATN, clinical, and cognitive measures a series of one-way analyses of variance (ANOVA) were run. Before running the ANOVA, we checked for the homoscedasticity of the data using the Levene’s test. For data with unequal variance, we used the Welch ANOVA which better controls for type I error in case of heterogeneity of variance.^33^

#### Comparing dynamic connectivity patterns between patient clusters

For the temporal CAPs metrics, a series of mixed ANOVA were run to investigate interaction effects between cluster assignment and specific metrics. CAP was inserted as within-factor, while patient cluster assignment was considered the between-factor. Each specific temporal metric represented the dependent variable and considered independently from the other metrics. A p-value<0.05 was set as significant. For this analysis we considered the full set of temporal metrics, although for the transition probabilities matrix only the diagonal was here considered. This decomposition enabled to retrieve a single value for each CAP, in line with the other metrics.

### Multivariate analysis of ATN and dynamic connectivity measures

Multivariate analyses were performed at two different levels, aimed at complementing the univariate findings described above. First, in each identified CAP we run a factorial analysis (varimax rotation) on the 7 temporal metrics. Factors were retained based on the eigenvalue silhouette and if explaining more than 10% of variance.^18^ Kaiser-Meyer-Olkin (KMO) test was run to test the sample size adequacy of the factorial analysis, by examining the proportion of variance among variables that might be common variance, i.e., that might be caused by underlying factors. More in details, high KMO values (>0.7) indicate that most of the partial correlations are small compared to the correlations; low values (<0.7) indicate that the partial correlations are relatively large, suggesting that the variables are not well suited for factor analysis. Latent factors for each CAP were then independently analyzed through a mixed ANOVA, with latent components as the within-factor, and patient cluster assignment as between-factor, according to the same criteria applied for the univariate analysis. The same factorial analysis was applied on the ATN outcomes to further reduce the dimensionality of these measures and improve the interpretability of results.

Finally, for each CAP, ATN measures and temporal metrics were fed into a canonical correlation analysis (CCA) to investigate the multivariate correlation patterns between ATN and dFC temporal features (see paragraph S1.4 in the **Supplementary Material** for details). Significant CAP-modes were entered into a one-way ANOVA to investigate differences among patient clusters.

#### CAP spatial analysis

We run a post-hoc spatial voxel-wise analysis to compare the subject-level CAP topographies across patient clusters. We applied a non-parametric approach suitable for high dimensional data (fsl-randomise; fsl.fmrib.ox.ac.uk/fsl/fslwiki/Randomise) at a threshold-free cluster enhancement (n=5000 permutations, p<0.05 FWE-corrected with a minimum size k=50). Significant results were registered to the fsaverage template and showed at surface level.

### Predicting patient clusters from CAPs temporal dynamics

In order to investigate the predictive power of dFC to predict patients’ classification computed with the ATN measures, we applied a random forest (RF) algorithm (paragraph S1.5 in the S**upplementary Material**), independently for each CAP. We used the latent factors from the factorial analysis on the temporal CAP metrics as predictors with a k-fold cross-validation (k=5) and a fixed random seed for each run. For the test set, balanced accuracy, and f1 score (weighted) of the CAPs-based RF classifiers were compared across CAPs to identify which CAP allowed to retrieve the most accurate patient classification based on the ATN patterns. A logistic regression analysis was implemented to assess the replication of the RF results.

## RESULTS

### Posterior cingulate cortex co-activation patterns

Consensus clustering of rs-fMRI PCC-activation frames in CU showed the optimal (stable) number of CAPs to be 5 (**Fig. S1**), in line with previous literature using PCC seed.^20^ CAP1 encompassed brain parcels belonging to the DMN mainly mapping to frontal and parietal regions, while regions in sensory regions showed co-deactivation. CAP2 included brain parcels overlapping with the visual, sensorimotor, and dorsal attention (DAN) networks. CAP3 overlapped mainly with the parietal and temporal regions of the DMN and co-deactivated in DAN regions. CAP4 showed an occipital-temporal gradient. Finally, CAP5 showed a widespread fronto-parieto-temporal pattern. These dynamic patterns were similar in the patient group (**Fig. 2**). High spatial correlation between patients and controls were reported for all the group-averaged CAPs (CAP1, r=0.982; CAP2, r=0.978; CAP3, r=0.980; CAP4, r=0.978; CAP5, r=0.973, with all p<0.00001), excluding potential biases in CAPs construction. When group averaged CAPs were projected at the cortical surface (parcel-level), a significant difference was observed in each CAP across the seven Yeo’s^34^ resting state network topographies (one-way ANOVA: p<0.0001 for all CAPs; **Fig. S2**).

**Figure 2.**
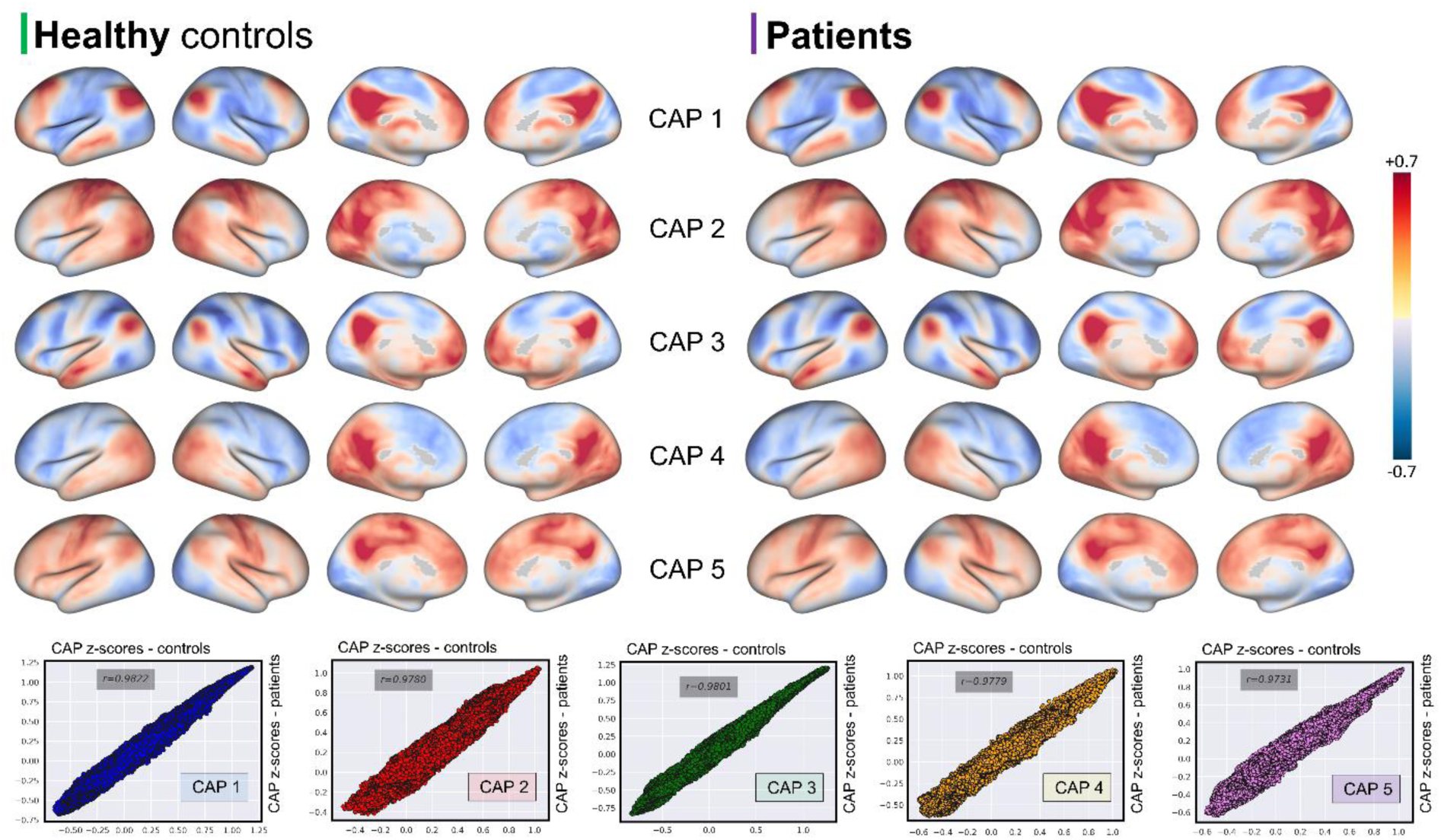
Posterior cingulate cortex CAPs in cognitively unimpaired controls and patients. CAPs for healthy controls (left panel) and patients within the clinical AD continuum (right panel) mapped onto the inflated cortical surface (32k vertices). Patients’ CAPs were obtained by averaging the PCC-activation frames assigned to one of the healthy control CAP (no clustering was run on patients rs-fMRI). High spatial correlations were reported for each CAP between groups (scatter plots and Pearson’s correlation (R) values are reported in the bottom panel), suggesting a high spatial stability of the PCC CAPs in the ageing and AD continuum.

### Data-driven patients’ stratification based on ATN measurements

Following UMAP dimensionality reduction, patient data were clustered into three groups (K-means algorithm), according to both silhouette and elbow methods (**Fig. S3**).

The three patient clusters represented well segregated groups in the UMAP space (**Fig. S4**, top panel), with dimensions equals to 45, 35, and 19 patients, respectively. While the main UMAP analysis was run with the default parameters, we also performed an additional sensitivity analysis with different UMAP parameters followed by K-means clustering (K=3) to assess the stability of the results. Group assignment accuracy between the main UMAP analysis and additional UMAP runs was very high (average: 0.952, range: 0.71–1.0), indicating that patients were assigned to the same clusters with very high probability at each UMAP-KMeans run (**Fig. S4**). Moreover, cluster sizes were comparable across UMAP-Kmeans runs indicating high results’ stability (**Fig. S4**).

### ATN profile across patients’ clusters

The three clusters were different for almost all the ATN measures (p<0.001)(**Table 2**). Welch’s ANOVA was used for Aβ and tau from CSF, whole brain composite amyloid-PET and tau-PET, hippocampal volume. For the remaining measures a classic one-way ANOVA was run. The *first* cluster showed the highest accumulation of A and T features compared to the other clusters. CSF tau, ptau and amyloid level were higher in the *first* cluster compared to both the *second* and *third* clusters (CSF-amyloid metrics were multiplied by -1 for consistencies with the other measures). Similar patterns were reported for whole brain composite amyloid- and tau-PET. Notably, the *third* cluster showed the highest rate of medial temporal atrophy and hypometabolism (N).

**Table 2.**
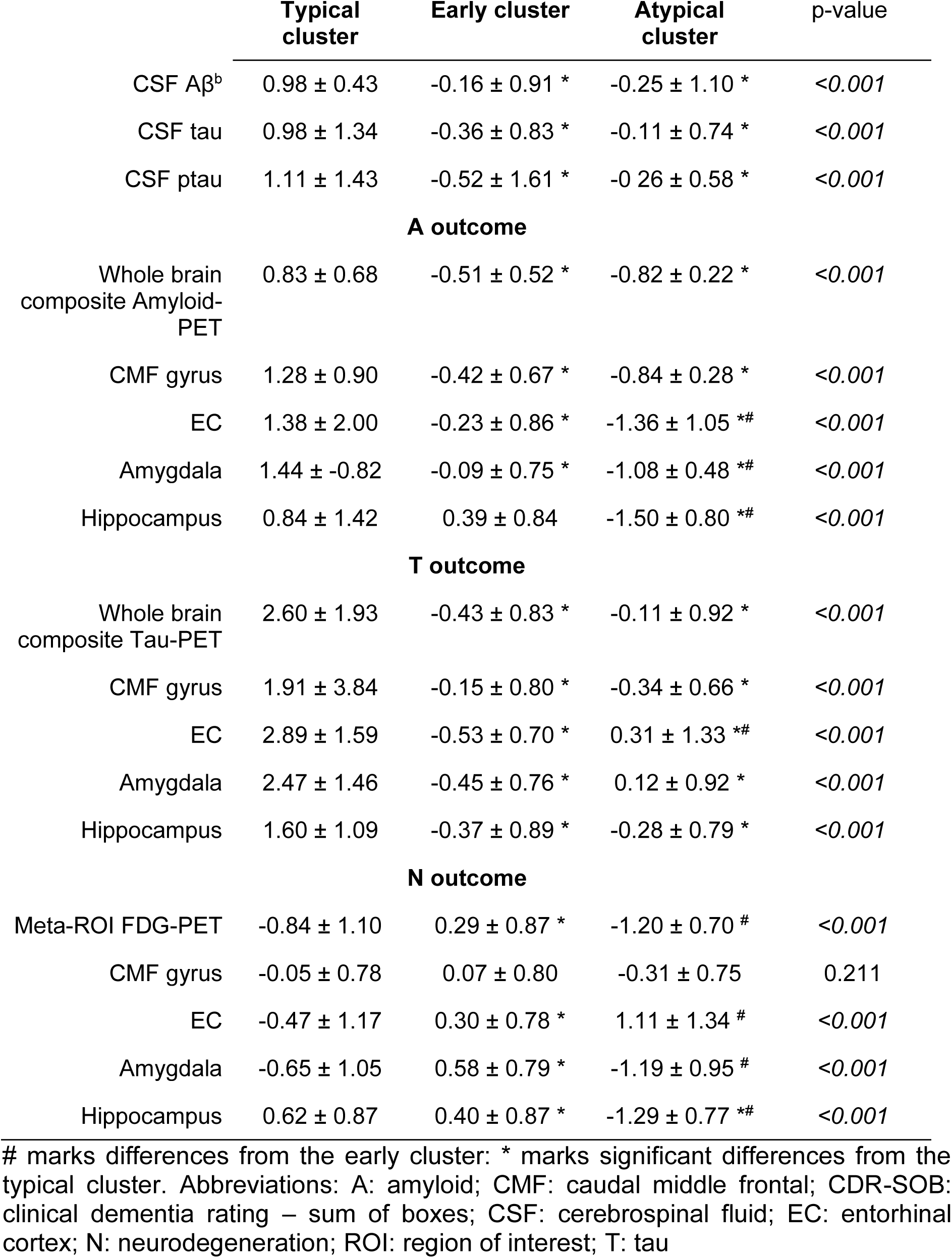
ATN profile of patients’ subgroups. Data are shown as z-score according to the healthy control sample included in the analysis. Aβ was inverted for congruency with the amyloid-PET measures (high values indicative of higher brain Aβ accumulation).

This finding was confirmed when ANOVA was performed considering the ATN latent factors from the factorial analysis. Three latent factors explained more than 50% of data variance (KMO=0.769), closely reflecting the ATN framework. The first factor loaded on T measures (both CSF and PET), the second factor loaded on A measures (CSF (multiplied by -1) and PET), while the third factor expressed neurodegeneration (N), loading on volumetric and metabolic measures (**Fig. 3**). All the three components were different among groups (Factor 1 (T): F=19.22, p<0.0001, np^2^=0.29; Factor 2 (A): F=33.73, p<0.0001, np^2^=0.41; Factor 3 (N): F=33.52, p<0.0001, np^2^=0.41). Post-hoc analysis showed that T accumulation was highest in the *first* cluster (vs both *early* and *atypical*; p<0.001) while being similar between the *second* and *third* (p=0.616)(**Fig. 4**, bottom panel). A gradient was found for A accumulation, with the highest value for the *first* cluster compared to both the *second* and *third* clusters (p<0.0001) and lower in the *third* compared to *second* (p<0.001). Regarding N, the *third* cluster showed the highest atrophy pattern (N) compared to both the *second* (p<0.0001) and *third* clusters (p=0.012), while the *first* cluster showed higher atrophy than the *second* one (p<0.0001).

**Figure 3.**
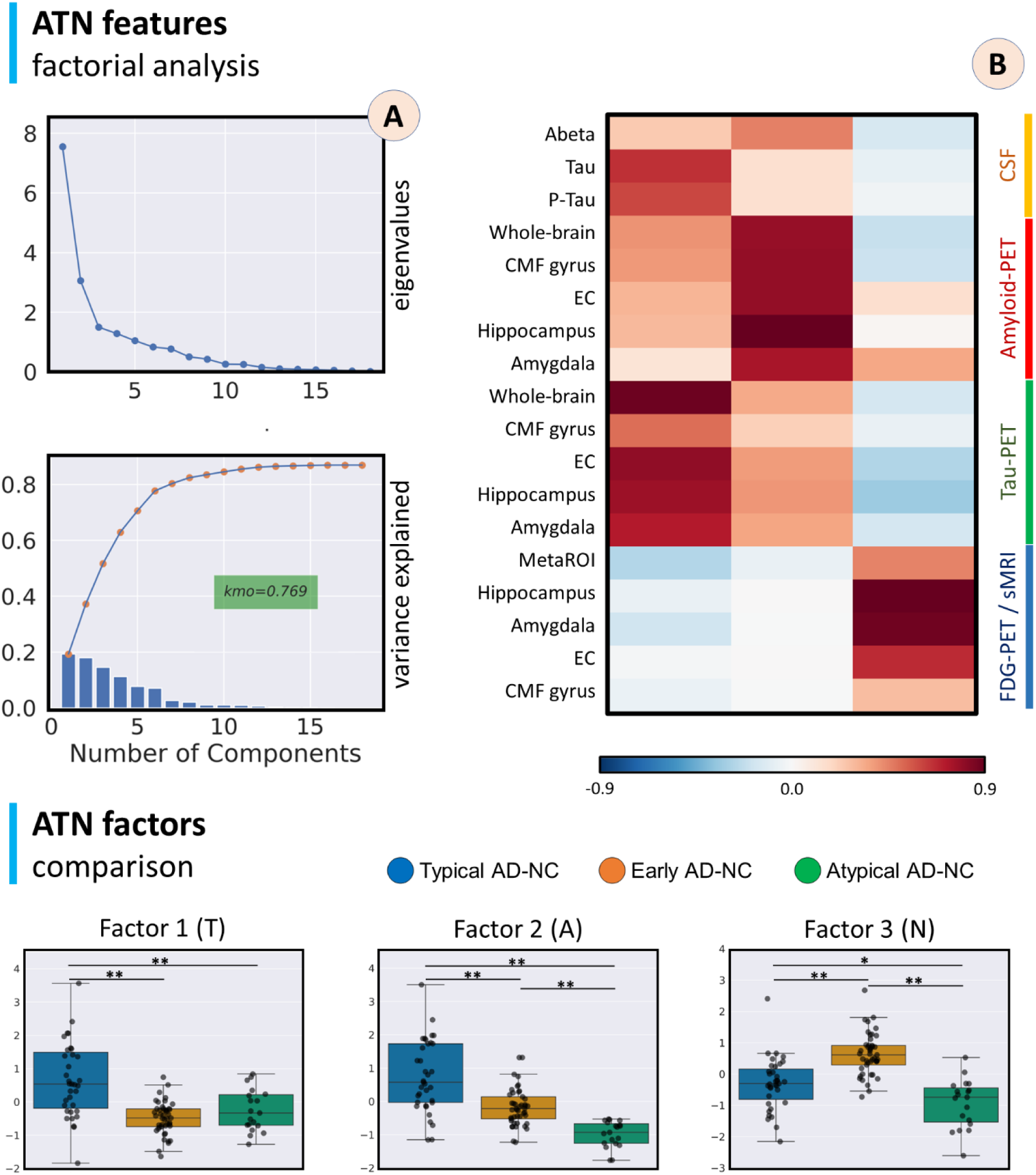
ATN measures factorial analysis. Top panels: factorial analysis of ATN measures in the ATN cohort. Amyloid, tau, structural, metabolism and CSF measures were z-scored according to mean and standard deviation of the control cohort before entering the factorial analysis. Panel A: eigenvalues and cumulative inter-individual variance explained by the latent factors. Panel B: loading matrix of the first three latent factors. On the right, different color lines are used to group the ATN variables (orange: CSF measures; red: amyloid-PET; green: tau-PET; blue: FDG-PET and brain volumetry from structural MRI (sMRI). Bottom panel: significant differences were reported between the three factors across the identified clusters. ** marks p < 0.001; * marks p<0.05.

**Figure 4.**
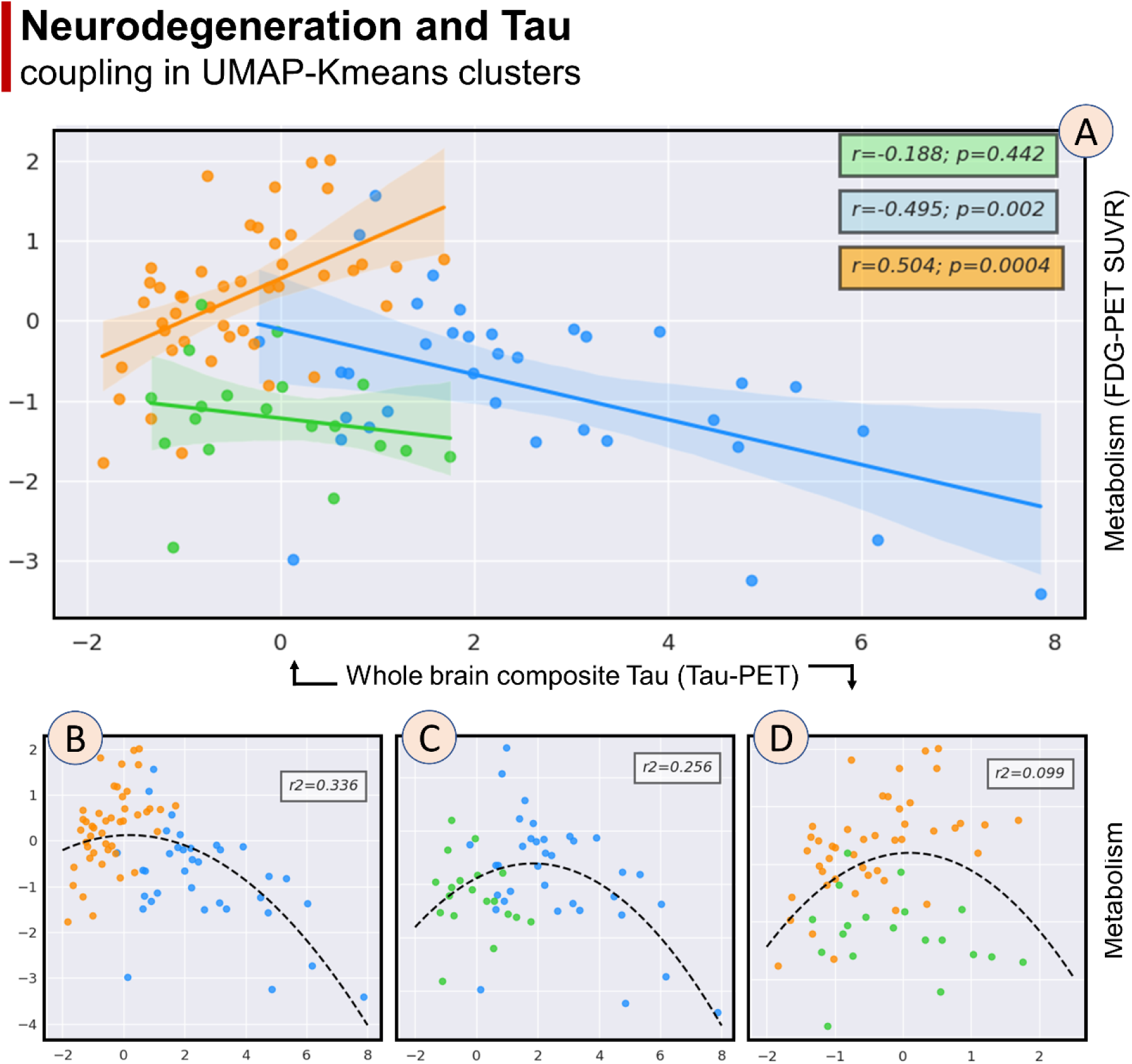
Metabolism and cerebral tau accumulation relation for patients’ clusters. Panel A: the relationship between neurodegeneration (’N’, assessed with FDG-PET MetaROI) and tau load (‘T’, assessed with whole brain composite tau-PET SUVR) showed typical trajectories in the early (orange) and typical (blue) cluster, while a third cluster (’atypical’; green) showed no relationship between T and N. When data were grouped two by two, the best polynomial fit (indicative of a U-shaped inversion typical of AD pathology) was reported when early and typical clusters were combined (R2=0.336; panel B). Lower values were reported when considering atypical clusters (panel C and panel D).

To further characterize these clusters, we explored the relationships between whole brain composite tau-PET SUVR and FDG-PET metabolism in these groups. Specifically, we employed a quadratic polynomial curve fitting with a degree of two for each pair of patient clusters, aimed at identifying non-linear relationship between these measures as suggested by previous studies.^35,36^ To evaluate the goodness of fit, we calculated the R^2^ value for each pair, which allowed us to determine the clusters that best represented, with a winner-take-all approach, the Tau-metabolism quadratic relationship. The three clusters showed peculiar relationships between these measures. The *first* cluster showed a strong negative correlation between metabolism and whole brain composite tau accumulation (r=-0.495, p=0.002; blue dots in **Fig 3**), while these measures were positively correlated in the *second* cluster (r=0.504; p=0.0004; orange dots in **Fig 3**). On the contrary, the *third* cluster showed no significant correlation between these outcomes (r=-0.188; p=0.442; green dots in **Fig 4**). In order to assess for the presence of an inverted U-shape relationship between metabolism and cerebral tau, previously described in AD^35,36^ and suggestive of an initial compensatory stage (high tau accumulation and high metabolism) followed by a detrimental stage (high tau and low metabolism), the different patient clusters were pooled together in groups of two. When performing a series of quadratic regressions, we found the largest predictive power by pooling the *first* and *second* clusters (R^2^=0.336). On the contrary, pooling the *first* and *third* clusters resulted in a drop of the quadratic predictive power (R^2^=0.256). Pooling the *second* and *third* clusters resulted in the smallest quadratic predictive power (R^2^=0.099). Overall, these results could suggest AD-NC (with different onset) for the *first* and *second* clusters, while the third one showed an atypical T-N trajectory (**Fig. 4**).

Based on these and above results, the *first* cluster could represent a subset of patients with a *typical* ATN profile in an advanced biological stage. The *second* cluster could represent an initial (*early*) stage of the typical ATN profile. Finally, the *third* cluster was suggestive of an *atypical* ATN pattern, highly suggestive of SNAP. A specific color code (orange for *early*, blue for *typical*, and green for *atypical*) was consistently used in all the figures throughout the manuscript to improve the readability.

### Sociodemographic, clinical, and cognitive features

The three patient clusters were comparable for gender distribution, education, and total intracranial volume (p=0.954, p=0.351, and p=0.203, respectively). Conversely, a significant age effect was reported (p=0.029). To account for possible confounding effects, age was linearly regressed from all the cognitive and clinical scores.

The three groups were different in terms of clinical severity and cognitive impairment (**Fig. 5**; **Table S1)**. A significant group effect was reported for MMSE score (Welch’s ANOVA F=18.48, p<0.001; np^2^=0.27), CDR-SOB (Welch’s ANOVA F=13.95, p<0.001; np^2^=0.22), ADAS-13 (one patient from the *atypical* cluster was removed due to missing value; Welch’s ANOVA F=20.54, p<0.001; np^2^=0.30), composite memory (ANOVA F=21.12, p<0.001; np^2^=0.31), composite executive functions (Welch’s ANOVA F=5.43, p=0.007, np^2^=0.10), and language abilities (Welch’s ANOVA F=5.61, p<0.001; np^2^=0.09). On the contrary, visuospatial abilities were not different between groups (ANOVA F=1.58, p=0.212; np^2^=0.03). In all significant ANOVAs, the post-hoc analysis showed a significant difference between the *early* group and the two other patients’ clusters for all the measures, suggesting a similar degree of cognitive impairment and clinical severity between *typical* and *atypical* clusters (**Fig. 5**). Results were similar considering data without regressing age.

**Figure 5.**
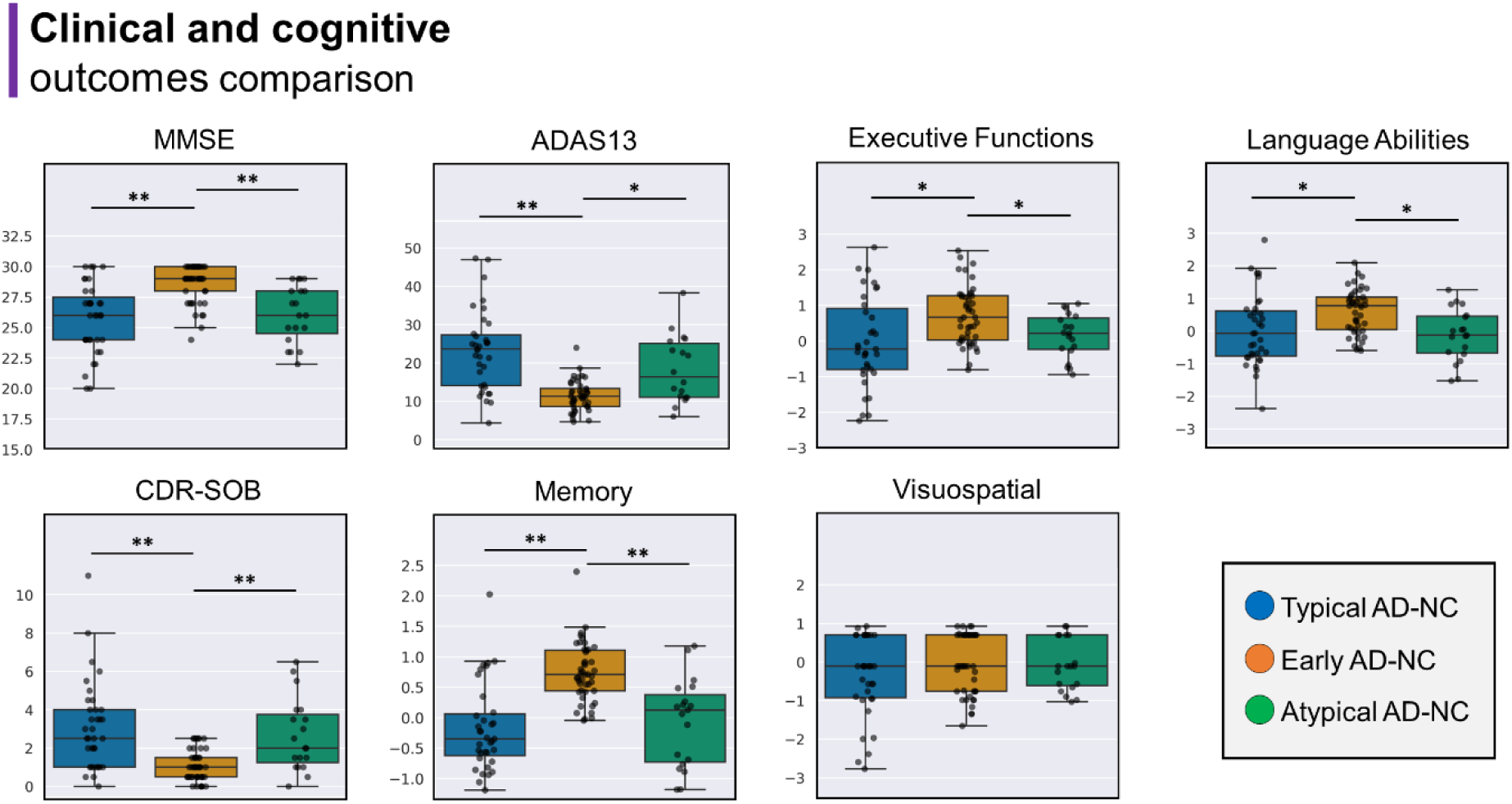
Clinical and cognitive comparison across patient clusters. Clinical and cognitive outcomes are reported as uncorrected data, while statistical analysis was performed on the age-corrected outcomes to account for potential age effects. ** marks p < 0.001; * marks p<0.05.

### CAPs across patients’ clusters

#### Univariate analysis

Temporal metrics were corrected for age effect in the same way reported for the cognitive/clinical outcomes. Among the temporal metrics included in the study, a significant group*CAP interaction effect was reported for average duration (F=2.059, p=0.039; np2=0.041), resilience (F=2.83, p=0.003; np2=0.056), subject entries (F=2.87, p=0.004; np2=0.056), counts (F=2.10, p=0.035; np2=0.042), and within-CAP transition probabilities (F=2.83, p=0.005; np2= 0.056). Betweenness (F=1.419, p=0.187), and out degree (F=0.86, p=0.553), showed no interaction effect (see paragraph S2.1 in the **Supplementary Material** and **Fig. S5** for post-hoc results).

#### Multivariate analysis

Five factorial analyses including the temporal metrics for each CAP independently were run. We found that the temporal metrics could be reduced to three main factors which together explained more than 80% of data variance in all the five CAPs. The first factor mainly loaded on the within-CAP transition probabilities, subject entries, resilience, and average duration, describing quantitative CAP patterns, explaining around the 40% of variance. The other two factors loaded on counts, out degree, and betweenness, indicative of CAP-dynamic features. This pattern was reported for all the CAPs. Similarly, the amount of variance explained by these three components was equivalent across CAPs (**Fig. S6**).

The mixed ANOVA run independently on each CAP and including the first three factors showed a significant group effect (F=4.70, p=0.011; np2= 0.084) for CAP1. Post-hoc analysis revealed significant differences between *typical* and *atypical* clusters, in line with the univariate analysis. Notably, CAP4 showed a significant group*factors interaction effect (F=6.33, p=0.002; np2=0.12). Post-hoc analysis revealed again a significant difference between *typical* and *atypical* for the first and second factors (p<0.05), while the third component showed a significant difference between *typical* and *early* clusters (p<0.05) suggestive of a specific temporal-CAP effect between these groups (**Fig. 6**, top-left panel).

**Figure 6.**
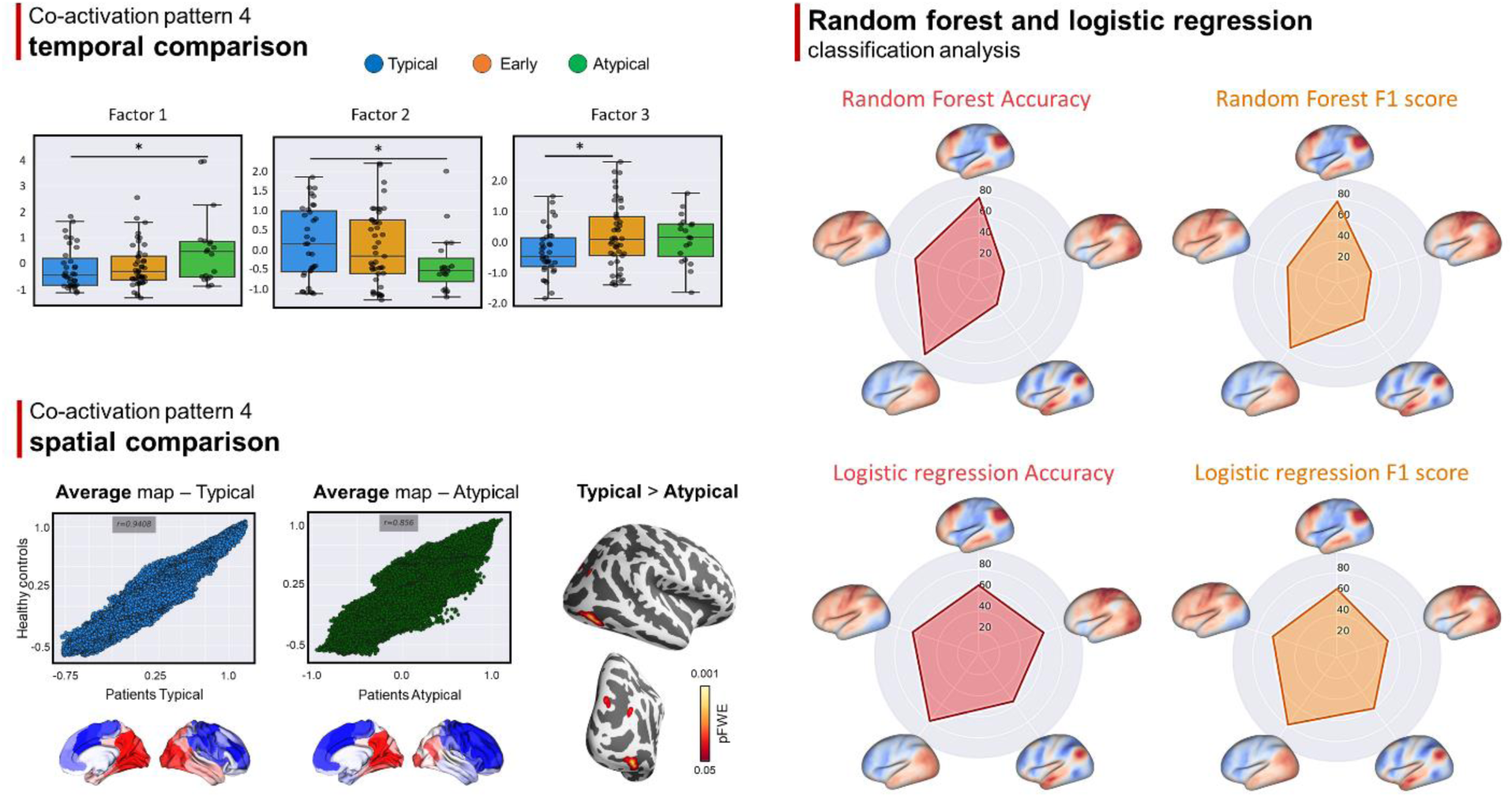
Dynamic connectivity differences across patient clusters. Left-top panel: significant differences in CAP4-temporal latent factors between patients’ clusters; each dot represents a patient; * marks significant post-hoc differences. Left-bottom panel: voxel-wise spatial differences within CAP4 between typical and atypical clusters (p<0.05, FWE corrected). Reduced similarity between group averaged CAP4 in the atypical group compared to controls is reported; each dot represents a voxel value from the group averaged CAP4 map. Right panel: Test set classification results applying a random forest classifier on CAPs temporal patterns (latent factors; see Fig. 4). CAP1 and CAP4 showed the highest performance in classifying typical and atypical patterns according to the UMAP-Kmeans algorithm applied to ATN measures (assessed with balanced accuracy and F1-scores). Result for CAP4 was confirmed by means of a linear logistic regression (both accuracy and f1 scores around 80%; all the other CAPs showed scores around 60%).

The CCA reinforced these results, with CAP1 and CAP4 being the most involved between patient clusters. The first mode of CAP1 exhibited significant group effects between the *typical* and *atypical* clusters. Similarly, the first mode of CAP4 displayed significant patterns and group effects between these two ATN clusters (Details are reported in the paragraph S2.2 in the **Supplementary Material** and **Fig. S7**).

### Predictive modeling using latent CAP factors

This analysis was run using as features the temporal CAP latent factors and limited to the classification of *typical* and *atypical* individuals, due to their similar cognitive impairment but significantly different ATN profile. In line with the multivariate findings, the three main latent factors from CAP1 and, especially, CAP4 showed the highest balanced accuracy when included in the RF classification algorithm (CAP1: 0.78; CAP4: 0.88) and f1 score (CAP1: 0.78; CAP4: 0.78) for predicting patient clusters derived from the ATN measures. On the contrary, the three latent factors from CAP2, CAP3, and CAP5 showed lower predictive power (accuracy < 0.64; f1 score < 0.54) (**Fig. 6**). CAP 4 results were confirmed by the logistic regression analysis (**Fig. 6**).

### Spatial CAPs topology in patients

Based on the temporal CAPs results, we performed a post-hoc analysis to investigate spatial differences between the two CAPs mainly involved in group discrimination, that is CAP1 and CAP4 between *atypical* and *typical* clusters. This analysis showed no significant differences between patient clusters for CAP1. Notably, CAP4 showed a significant spatial difference between the *typical* and *atypical* clusters, in line with previous results on dynamical patterns (**Fig. 6**).

## DISCUSSION

The main novelty of this study lies in exploring the relationship between dFC and biological patterns in clinical MCI/AD patients stratified according to a data-driven approach. In line with our assumptions, we identified a group with a peculiar ATN pattern distinguished by a divergent pattern of tau/amyloid accumulation and neurodegeneration compared to the typical groups. Interestingly, when considering the PCC-CAPs, the analyses consistently revealed different dFC patterns, confirming that different ATN profiles also share selective dFC changes. Such dFC patterns were also highly accurate in differentiating patients with typical and atypical ATN profiles.

The application of dFC in AD research has been relatively limited compared to static FC.^30^ However, a few studies have emerged that utilize dynamic connectivity analyses, specifically employing a sliding windows approach, which have reported alterations in the temporal stability of connectivity in AD compared to healthy controls.^37–39^ Over a decade ago, Chang and Glover demonstrated that the coherence and phase of FC between the PCC and the rest of the DMN exhibit temporal variation.^40^ Further, clustering analysis has revealed that brain networks display dynamic yet topographically stable connectivity patterns that differ significantly from the time-average connectivity pattern.^41^ Building upon these previous observations, we expanded the scope by employing a PCC-CAPs approach. This methodology has the potential to identify both co-active and co-deactive states, which allows to investigate whether neurological disorders are associated with alterations in correlated and/or anti-correlated connectivity patterns that are linked to the clinical phenotype of the disease.^42,43^ Accordingly, we reported five distinct PCC-CAPs, with a consistent spatial topography pattern between controls and patients. These findings align with a previous study which utilized PCC for CAP computation and reported five main CAPs in a healthy cohort,^20^ many of which overlapped with the CAPs identified in our study. Two previous studies from the same research group analyzed PCC-CAPs in a sample of patients with disorders of consciousness.^24,44^ While these studies extracted a fixed number of eight CAPs, our study determined the number of CAPs based on cluster stability. Despite this difference, there are notable similarities between the CAPs, such as the presence of cortical and temporal DMN CAPs, as well as the visual-temporal CAP, which suggests the overall stability of this methodology,^44^ supporting the translational value in the AD context. In the context of AD, Adhikari et al. applied CAP in rs-fMRI signals in old TG2576 mice (animal model of amyloidosis). They reported CAP dynamics’ differences between diseased and wild animals,^45^ suggesting that CAPs could be used as predictors distinguishing between different molecular processes.

In this study, the CAP framework was complemented with a dual data-driven strategy (low-dimensionality and clustering) on ATN measures to identify latent groups within the clinical AD continuum. We identified three patient clusters based on the ATN profile, providing new insights into the underlying biological processes. Two of these clusters aligned with the well-established pathophysiological distribution of the ATN measures, suggestive of different timelines of the pathological changes. A cluster exhibited characteristics indicative of an early pathophysiological stage of AD, in line with the subtle cognitive/clinical impairment. The second cluster demonstrated a clinical/cognitive profile consistent with the manifest stage of the AD cascade, marked by high levels of both Aβ and tau pathology, as well as widespread neurodegeneration. Interestingly, our analysis revealed a third cluster displaying an *atypical* ATN profile, suggestive of a different underlying syndrome. Although presenting with clinical and cognitive manifestations associated with AD (as compared with the second cluster), these patients exhibited distinct biological features. Specifically, it exhibited the most pronounced pattern of neurodegeneration while displaying the lowest pattern of tau and amyloid accumulation, suggesting a limited tau pathology compared to the other clusters.

This *atypical* profile could be suggestive, at least partially, of the SNAP construct. Originally, SNAP included patients with an A-N+ profile,^46^ and subsequently it was extended to include patients with an A−T−N+ profile,^47^ which is in line with the data-driven results of this third cluster. SNAP is a heterogeneous biomarker-based concept that can represent different pathologies, such as primary age-related tauopathy, Lewy body disease, LATE, or argyrophilic grain disease.^11^ On the contrary, our connectivity analysis pointed to specific dFC alterations within this group compared to the *typical* cluster, rather than showing widespread alterations indicative of a lack of distinct connectivity changes. Indeed, while *typical* and *atypical* groups showed similar level of cognitive and clinical impairment, differences in temporal dFC were reported, involving CAP1 (cortical DMN co-activation pattern) and, mainly, CAP4 (occipito-temporal co-activation pattern with the PCC). A large body of literature suggests that DMN alterations are linked with the biology of AD.^13–15^ CAP1 dynamic features could distinguish between typical and atypical clusters with high accuracy (around 80%), suggesting different DMN alterations in patients with comparable clinical impairment but distinct ATN profiles. This pattern was confirmed by the CCA analysis. The classification accuracy for distinguishing between typical and atypical cases was higher (approximately 90%) when employing temporal metrics derived from CAP4. CAP4 involves co-activation of the PCC within an occipital-parieto-temporal axis. Notably, occipital regions tend to remain unaffected in AD patients, even in the advanced stages of the disease,^48^ which implies that this CAP may play a role in non-AD neurodegenerative conditions. The CCA confirmed a connection between CAP4 and atypical ATN patterns. The reduction in spatial similarity observed between the group-averaged CAP4 of *atypical* cases compared to controls and *typical* clusters further reinforces the findings that CAP4 could serve as a distinctive ’fingerprint’ for these *atypical* ATN patients. Overall, these results suggest that PCC may serve as a central hub for both typical and atypical ATN patterns. However, distinct dynamic patterns likely underlie and differentiate specific profiles within this framework.

The results of our data-driven analysis, which identified three stable groups of patients, suggest that MCI/AD patients with non-AD-NC tend to cluster around a single group that may be more homogeneous than one might assume. This homogeneity is indicated not only by the robust identification of three main clusters but also by the dFC results highlighting specific differences. This raises questions about the nature of this atypical group. Although highly speculative, the atypical profile might resemble LATE-NC, characterized by memory impairment and medial temporal lobe degeneration without significant tau pathology.^26^ The advanced age of the patients within this cluster might support this speculation, as LATE-NC is often observed in older individuals.^26,49^ Recently, Tazwar et al.^50^ demonstrated lower transverse relaxation rate by combining ex-vivo MRI and histopathology, in a spatial pattern mainly involving the occipito-temporal cortex in LATE, in line with our dFC results. Moreover, Young and colleagues in a preliminary study investigated pathological progression patterns of TDP-43 in several neurodegenerative diseases reporting a subtype labelled ‘corticolimbic predominant’ with greater TDP-43 deposition in temporal regions and the occipital cortex.^51^ To validate these findings, additional studies incorporating histopathology are necessary to ascertain whether atypical patients represent a well-defined pathophysiological condition or whether dFC alterations could represent a generic feature for distinguishing ATN typical and atypical patients. Indeed, additional pathophysiological features can be linked with this atypical cluster, such as vascular abnormalities or comorbidity with different pathological conditions, such as α-synuclein. Landau et al. assessed cognitively impaired patients categorized into A+T+ or A+T-. Their findings suggested a relation between α-synuclein and poorer cognition, particularly in cases when tau levels was low.^52^

This study has both limitations and strengths. One notable strength is that it applies a data-driven methodology to an extensive patient database within the clinical AD continuum, incorporating ATN and rs-fMRI data. However, it is important to acknowledge that our results’ interpretation regarding the atypical cluster remain speculative due to the absence of histological data. To address this gap in knowledge, future investigations should aim to examine whether the observed dFC alterations in the time-varying domain of the PCC align with specific histopathological findings. The length of ADNI rsfMRI data time series (around 200 volumes), although in line with most clinical studies, might not be optimal and further studies are needed to evaluate the stability of the reported CAPs. Finally, in this study we limited the analysis to the PCC, due to its vulnerability in AD,^30^ and its role as main connectivity hub of the brain^53^, while other regions could show similar divergent patterns within the ATN continuum.

In conclusion, by examining the relationships between ATN measures and dFC patterns, we can gain insights into the mechanisms driving disease progression and potentially identify biomarkers that differentiate typical and atypical clinical AD patients. We also suggest that dFC might be useful to characterize patients in the AD continuum. By means of functional connectivity (F), a new ATN(F) framework could help to unravel patients with a clinical manifestation of AD linked with non-AD pathological pathways.

### Ethics approval and consent to participate

All data included in this article were collected for research use as part of the Alzheimer’s Disease Neuroimaging Initiative (ADNI) project. Ethics approval for data collection in ADNI was obtained by each ADNI participating institution’s institutional review board. All participants gave written informed consent at participating institutions. The authors of this paper were granted approved access to the ADNI data, and the ADNI Data Sharing and Publications Committee (DPC) approved this paper for submission.

## Consent for publication

Not Applicable

## Funding

MC was supported by Fondazione Cassa di Risparmio di Padova e Rovigo (CARIPARO)— Ricerca Scientifica di Eccellenza 2018 (Grant Agreement number 55403); Italian Ministero della Salute, Brain connectivity measured with high-density electroencephalography: a novel neurodiagnostic tool for stroke (NEUROCONN; RF-2018-1236689); Celeghin Foundation Padova (CUP C94I20000420007); BIAL foundation grant (No. 361/18); Horizon 2020 European School of Network Neuroscience— European School of Network Neuroscience (euSNN), H2020-SC5-2019-2 (Grant Agreement number 860563); Horizon 2020 research and innovation program; Visionary Nature Based Actions For Heath, Wellbeing & Resilience in Cities (VARCITIES), Horizon 2020-SC5-2019-2 (Grant Agreement number 869505); Italian Ministero della Salute: Eye-movement dynamics during free viewing as biomarker for assessment of visuospatial functions and for closed-loop rehabilitation in stroke (EYEMOVINSTROKE; RF-2019-12369300). IBG, LB and GM acknowledge support from Fondazione CariVerona (Bando Ricerca Scientifica di Eccellenza 2018, EDIPO project, num. 2018.0855.2019) and MIUR D.M. 737/2021 “AI4Health: empowering neurosciences with eXplainable AI methods”. Data collection and sharing for this project was funded by the Alzheimer’s Disease Neuroimaging Initiative (ADNI) (National Institutes of Health Grant U01 AG024904) and DOD ADNI (Department of Defense award number W81XWH-12-2-0012). ADNI is funded by the National Institute on Aging, the National Institute of Biomedical Imaging and Bioengineering, and through generous contributions from the following: AbbVie, Alzheimer’s Association; Alzheimer’s Drug Discovery Foundation; Araclon Biotech; BioClinica, Inc.; Biogen; Bristol-Myers Squibb Company; CereSpir, Inc.; Cogstate; Eisai Inc.; Elan Pharmaceuticals, Inc.; Eli Lilly and Company; EuroImmun; F. Hoffmann-La Roche Ltd and its affiliated company Genentech, Inc.; Fujirebio; GE Healthcare; IXICO Ltd.; Janssen Alzheimer Immunotherapy Research & Development, LLC.; Johnson & Johnson Pharmaceutical Research & Development LLC.; Lumosity; Lundbeck; Merck & Co., Inc.; Meso Scale Diagnostics, LLC.; NeuroRx Research; Neurotrack Technologies; Novartis Pharmaceuticals Corporation; Pfizer Inc.; Piramal Imaging; Servier; Takeda Pharmaceutical Company; and Transition Therapeutics. The Canadian Institutes of Health Research is providing funds to support ADNI clinical sites in Canada. Private sector contributions are facilitated by the Foundation for the National Institutes of Health (www.fnih.org). The grantee organization is the Northern California Institute for Research and Education, and the study is coordinated by the Alzheimer’s Therapeutic Research Institute at the University of Southern California. ADNI data are disseminated by the Laboratory for Neuro Imaging at the University of Southern California.

## Abbreviations

A: amyloid
ADNI: Alzheimer’s Disease Neuroimaging Initiative
CAPs: co-activation patterns
dFC: dynamic FC
N: neurodegeneration
PCC: posterior cingulate cortex
PET: positron emission tomography
rs-fMRI: resting state functional magnetic resonance imaging
T: tau

## Conflict of interest

none

## Data Availability Statement

All data used in this manuscript are available to the public at the ADNI data repository at the Laboratory of Neuroimaging (http://adni.loni.usc.edu). Derived data are available from the authors upon request.

## Acknowledgements

Not Applicable

## Notes

### Competing Interest Statement

The authors have declared no competing interest.

### Funding Statement

This study did not receive any funding.

### Author Declarations

All data included in this article were collected for research use as part of the Alzheimer's Disease Neuroimaging Initiative (ADNI) project. Ethics approval for data collection in ADNI was obtained by each ADNI participating institution's institutional review board. The authors of this paper were granted approved access to the ADNI data, and the ADNI Data Sharing and Publications Committee (DPC) approved this paper for submission.

