## Supplementary Material for "Functional dynamic network connectivity differentiates biological patterns in the Alzheimer’s disease continuum"

**This PDF file includes:**

Supporting text
Figures S1 to S7
Tables S1

#### S1. SUPPLEMENTARY METHODS

*S1.1 Resting-state fMRI parameters and preprocessing*

For resting-state fMRI the following sequence parameters were used: repetition/echo time (TR/TE)=3000/30 ms, FA=90°, FOV=220x220x163 mm, 3.4-mm isotropic voxel size. 200 fMRI volumes were acquired in almost all subjects, with minimal variations in a small subset (n=197). Details are reported in https://adni.loni.usc.edu/adni-3/procedure-manuals/. Minimal preprocessing was firstly applied to the data including removal of the first 5 volumes, interleaved slice-timing correction, motion realignment (MCFLIRT), and 4D mean intensity normalization. The six motion parameters (plus their derivatives), mean white matter (WM)/CSF signals and a linear trend component were then linearly regressed out from the minimally preprocessed data. WM and CSF mean signals were extracted from the corresponding partial volume maps derived from the tissue segmentation of the T1-w scans and registered to the rs-fMRI native space using the Boundary-Based Registration algorithm (6 DOF), after erosion and binarization with a threshold of 0.8. The residuals resulting from this analysis were subsequently band-pass filtered (0.01-0.08 Hz). Finally, the preprocessed rs-fMRI volumes were spatially normalized to the 2-mm MNI space using a linear and non-linear registration (FLIRT and FNIRT) and smoothed with a 6-mm FWHM kernel.

*S1.2 CAPs temporal metrics*

The following temporal metrics were analyzed: i) resilience: the likelihood of the signal to stay in a CAP K through time; ii) betweenness centrality: the number of times a node lies on the shortest path between other nodes; iii) average duration: average of the duration for which a CAP is sustained when entered; iv) subject entries: subject-specific counts of expressed CAPs; v) counts: number of frames; vi) out degree: the likelihood to exit a CAP towards any other; vii) transition probabilities: the probability for CAP i to directly enter CAP j, resulting in an asymmetric n×n matrix where n=number of CAPs.

*S1.3 UMAP algorithm*

UMAP, which adheres to the principles of Riemannian geometry and algebraic topology, represents the underlying structure of the input data. By relying on a graph layout algorithm, UMAP firstly builds a high dimensional graph representation of the data and then optimizes a low-dimensional graph to be as structurally similar as possible. It creates the so-called "fuzzy simplicial complex", a representation of a weighted graph, with edge weights representing the likelihood that two points are connected. To determine connectedness, UMAP extends a radius outward from each point, connecting points when those radii overlap. UMAP parameters were set to default values (i.e., an approximation of 15 neighbors and a minimum 0.1 Euclidean distance to obtain a two-dimensional embedding), in line with previous studies applying this algorithm.30 After reduction of the ATN features in the UMAP space, k-means was run to identify different clusters of patients. The optimal K was identified by applying the elbow and silhouette methods (ranging from 2 to 10). Finally, we performed a sensitivity analysis to ensure the robustness of these clusters. The k-means algorithm (maintaining K fixed to 3) was iteratively run using different combinations of UMAP parameters (neighbours=10, 15, 20, 25; minimum distance = 0.1, 0.25, 0.5; component = two- and three-dimensional). Clusters identified with this iterative procedure were compared in terms of accuracy using the standard assignment (i.e., k-means following standard UMAP parameters) as reference.

*S1.4 Canonical correlation analysis*

A permutation approach (n=1000) was applied to investigate the significance of the retrieved modes in contrast to a random distribution of the same data. CCA was applied to the whole ATN cohort, while modes significance was assessed separately for each AD clusters (identified with the UMAP-Kmeans approach), to take in account potential differences in the association between different ATN patterns, as highlighted in paragraph 3.3 from the main text. Specifically, we computed the r-value between modes for each cluster, and the largest group-value was compared with the null distribution to assess the significance of the CCA. Significance was set to a two-tailed p-value<0.05.

*S1.5 Random Forest algorithm*

For the random forest algorithm (a non-linear ensemble learning method that operates by constructing a multitude of decision trees) data were split into test and train data with a 25% and 75% proportion, respectively. The training dataset was used to identify the best set of hyperparameters, that is: estimators (from 200 to 2000 with step of 200), depth (from 1 to 10 with step of 1), sample split (2, 5, and 10), samples leaf (1, 2, and 4), and bootstrap (true or false). The best set of parameters identified with a n=5 fold cross-validation on the train set was as follows: max depth=9, min samples leaf=4, min samples split=5, and bootstrap=True.

#### S2. SUPPLEMENTARY RESULTS

*S2.1 CAPs across patients' clusters*

CAP1 was significantly different between *typical* and *atypical* clusters ($p<0.05$) for all the metrics included. Similarly, CAP3 showed significant differences mainly between *typical* and *atypical* clusters (resilience, subject entries, and transition probabilities), with an additional effect between *typical* and *early* clusters for subject entries. CAP4 showed a significant average duration difference between *typical* with both *early* and *atypical* clusters. Finally, CAP5 showed significant differences between *typical* and *early* clusters for the following metrics: counts, subject entries, resilience, and transition probabilities. The two latter metrics were significantly different also between *typical* and *atypical*. CAP2 showed no significant differences between patients' groups. Results are reported in Figure S5.

*S2.2 Different modes from canonical correlation analysis between ATN clusters*

For CAP1, the first two modes, compared to a random distribution, showed a statistically significant pattern (rmode1=0.597; ppermutation1=0.003; rmode2=0.571; ppermutation2<0.001). The ANOVA on the CAP1 modes showed a significant group effect for the first mode ($F=3.55$; $p=0.032$). In line with the univariate and factorial analysis, post-hoc analysis highlighted a significant difference between typical and atypical clusters. For the second mode, despite its significance, no group effects were reported on the one-way ANOVA ($p=0.768$). When considering the loadings for the first mode, a positive association between average duration and tau, mainly from CSF was found, as well as between subject entries/betweenness/counts and neurodegenerative patterns (though with negative loadings in this second case), suggesting a gradient along a more segregated dFC pattern linked with tau accumulation and atrophy (Fig. 6, top-right panel). CAP4 showed a significant pattern for the first mode (rmode1=0.646; ppermutation1=0.038). Notably, a significant group effect was reported ($F=5.44$; $p=0.006$), involving again typical and atypical clusters (post-hoc analysis). Positive loadings for the first mode encompassed average duration, subject entries, and counts and were positively associated with loading on the Aβ from CSF. The remaining ATN loadings were mostly negative, mapping especially on tau and amyloid-PET measures. This pattern suggests an atypical pattern between Aβ CSF (data from CSF were inverted to be consistent with PET data) and brain A/T coupled with a more segregated CAP4 dynamic.

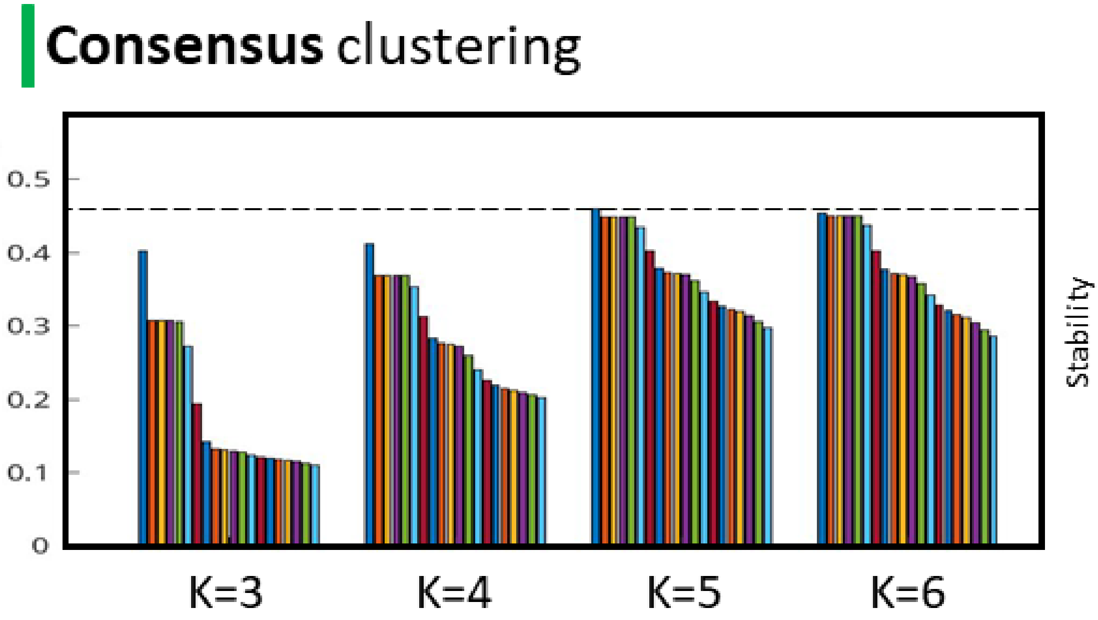


**Figure S1. Consensus clustering.**
The consensus clustering identified k=5 as the most stable set of clusters for the posterior cingulate cortex seed.

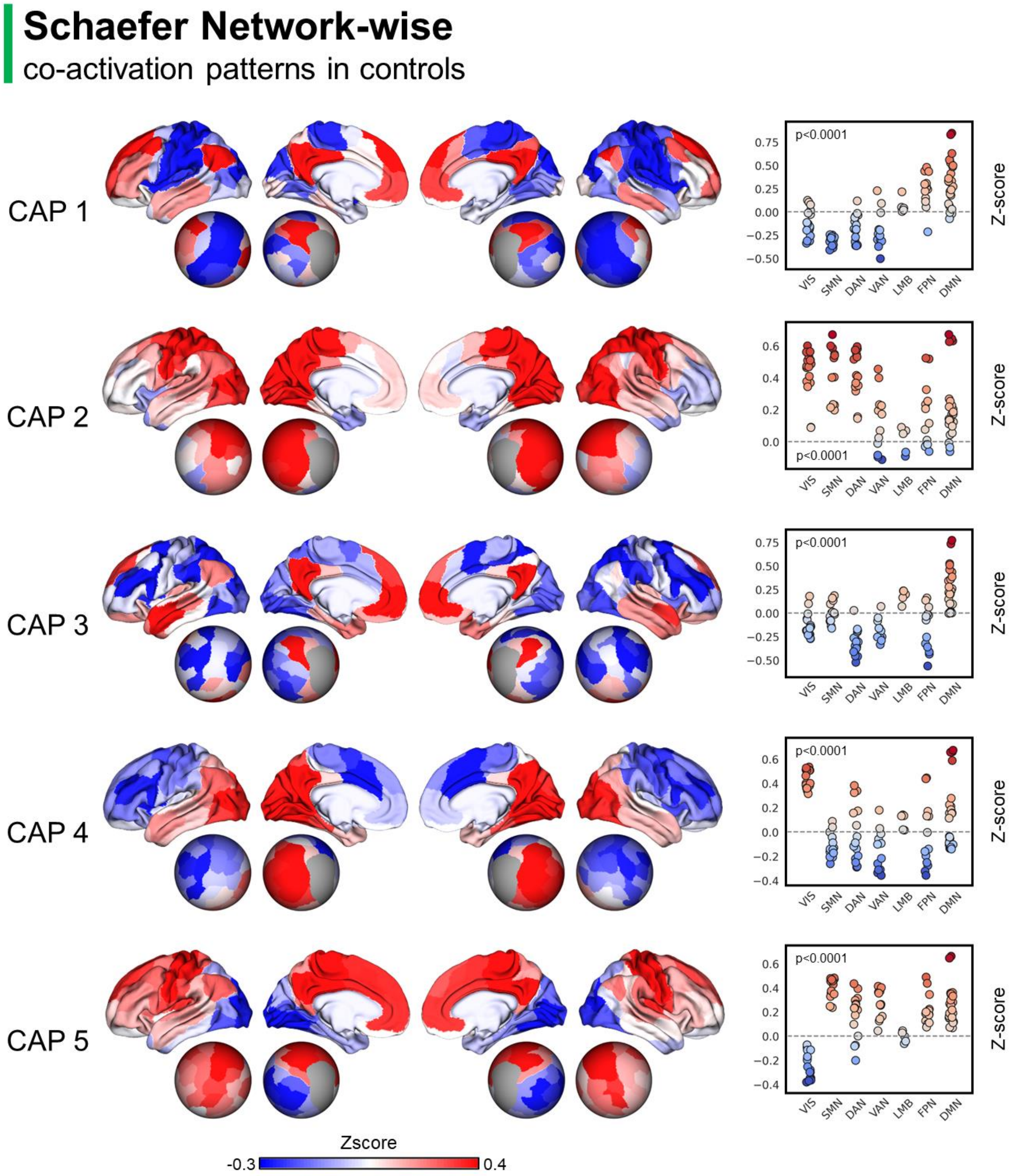


**Figure S2. Co-activation patterns identified healthy controls.**

Maps were projected at cortical surface level to the Conte69 brain template. Each cortical vertex was assigned to a specific parcel from the Schaefer 100 parcellation, and CAP z-score values were averaged within each parcel (cortical plots on the left). Parcels are also plotted at the spherical and flattened surfaces to improve visualization. A second stratification was computed according to Yeo's 7 resting state network parcellation (scatter plots on the right). Significant differences were reported for each CAP with respect to resting state networks by means of a one-way ANOVA on z-score values. P-values report the ANOVA main results.

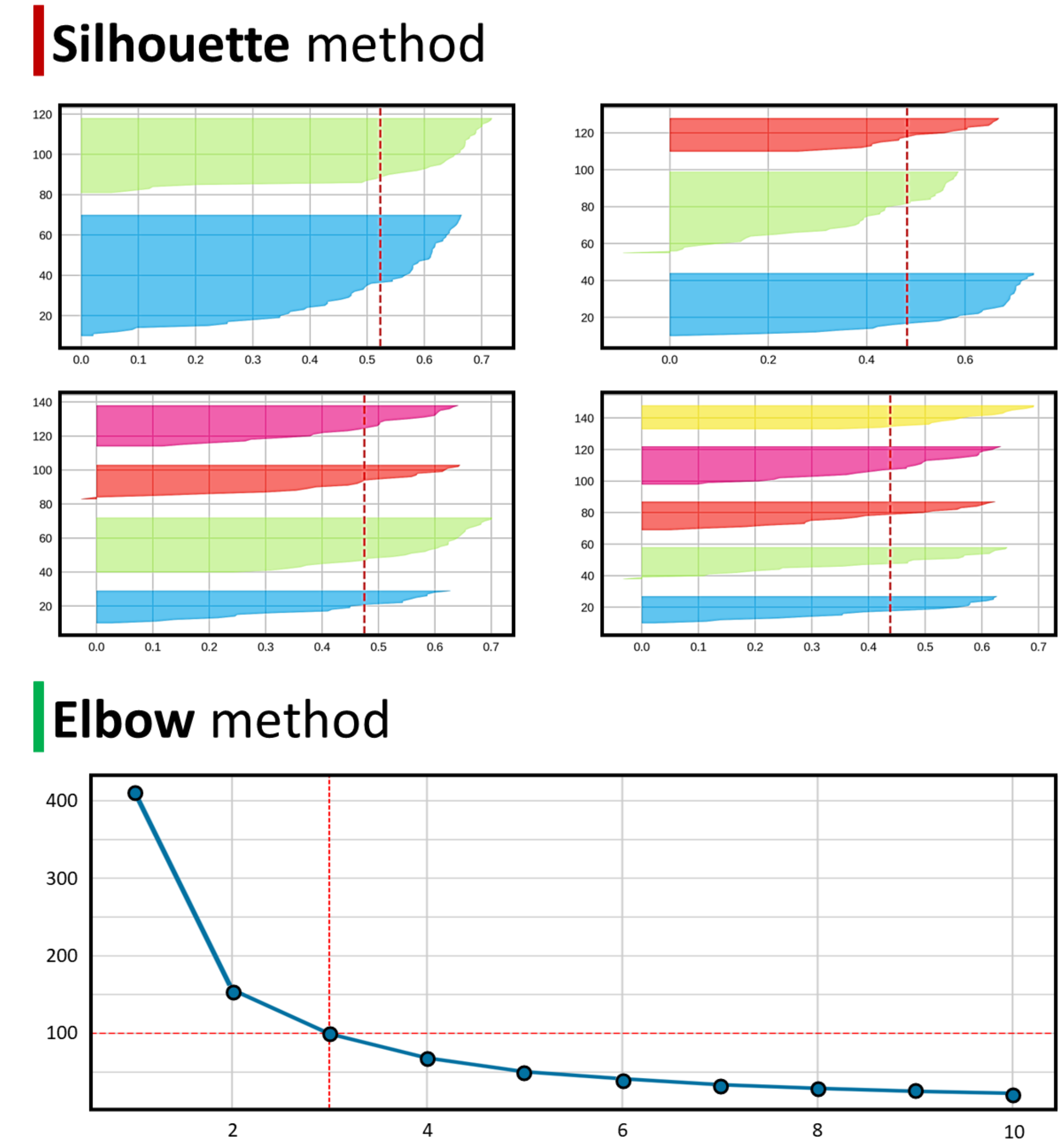


**Figure S3. Elbow and Silhouette method for UMAP-Kmeans.**
A number of cluster K=3 showed the most stable results, either using the silhouette (K from 2 to 5 are reported) or the elbow method.

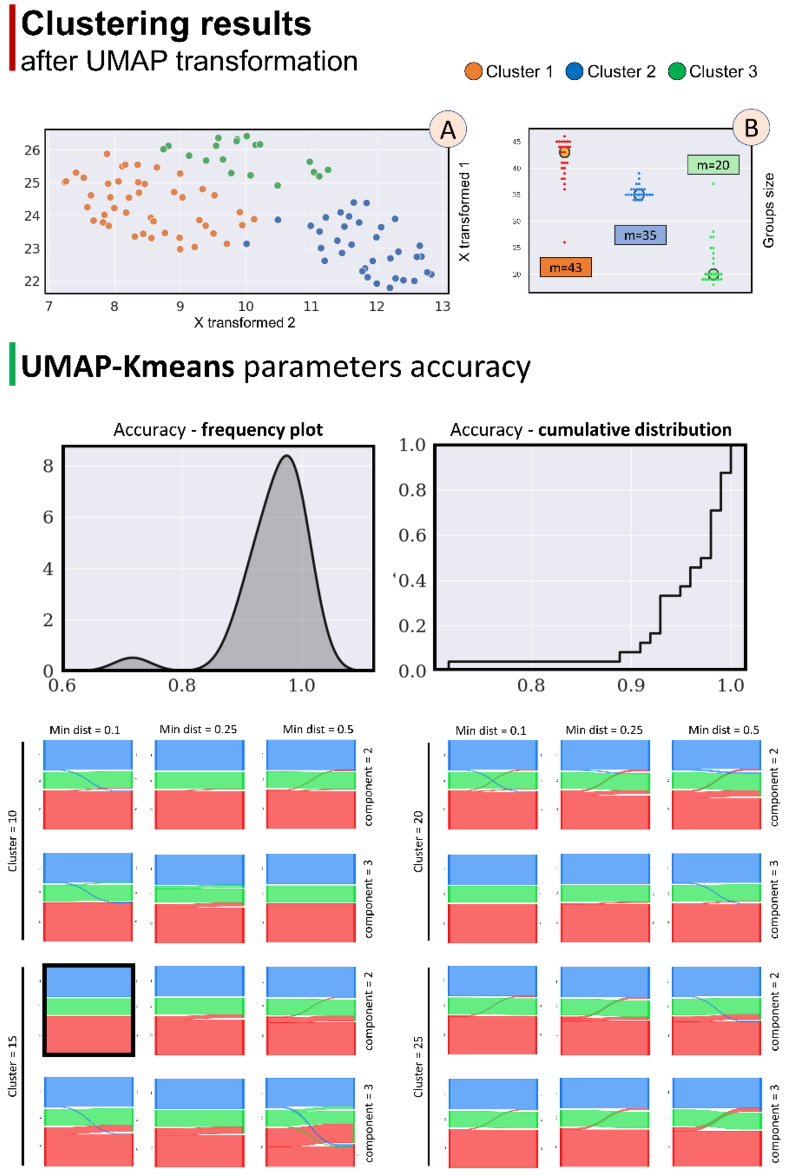


**Figure S4. UMAP sensitivity analysis.**

**Top panel (A):** we identified three groups of patients in the UMAP space based on 18 ATN features and using K-means clustering. Each dot represents a patient coloured according to the assigned cluster: orange=cluster 1; blue=cluster 2; green=cluster 3. **Top panel (B):** these three groups showed high stability across different UMAP parameters (m=median of the sample size across all n=24 UMAP-KMeans (K=3) runs). **Bottom panel**: UMAP results using different combinations of the following hyperparameters: minimum distance, components, and cluster. Top panel: Each UMAP-Kmeans run attributing a specific patient to a particular cluster was compared to the main UMAP analysis using the default parameters ("ground-truth"). Accuracy for patients' cluster attribution showed high values, ranging from 0.71 to 1.0, with an average value of 0.95 (left panel). To improve the visualization accuracy scores were plotted as cumulative distribution function (right panel). Bottom Panel: systematic comparisons between each cluster assignment for the different combinations of UMAP parameters. On the left of each panel is represented the distribution of patients within the three groups using the default parameters (highlighted in the black box), while on the right is represented patients' assignment using a different combination of parameters.

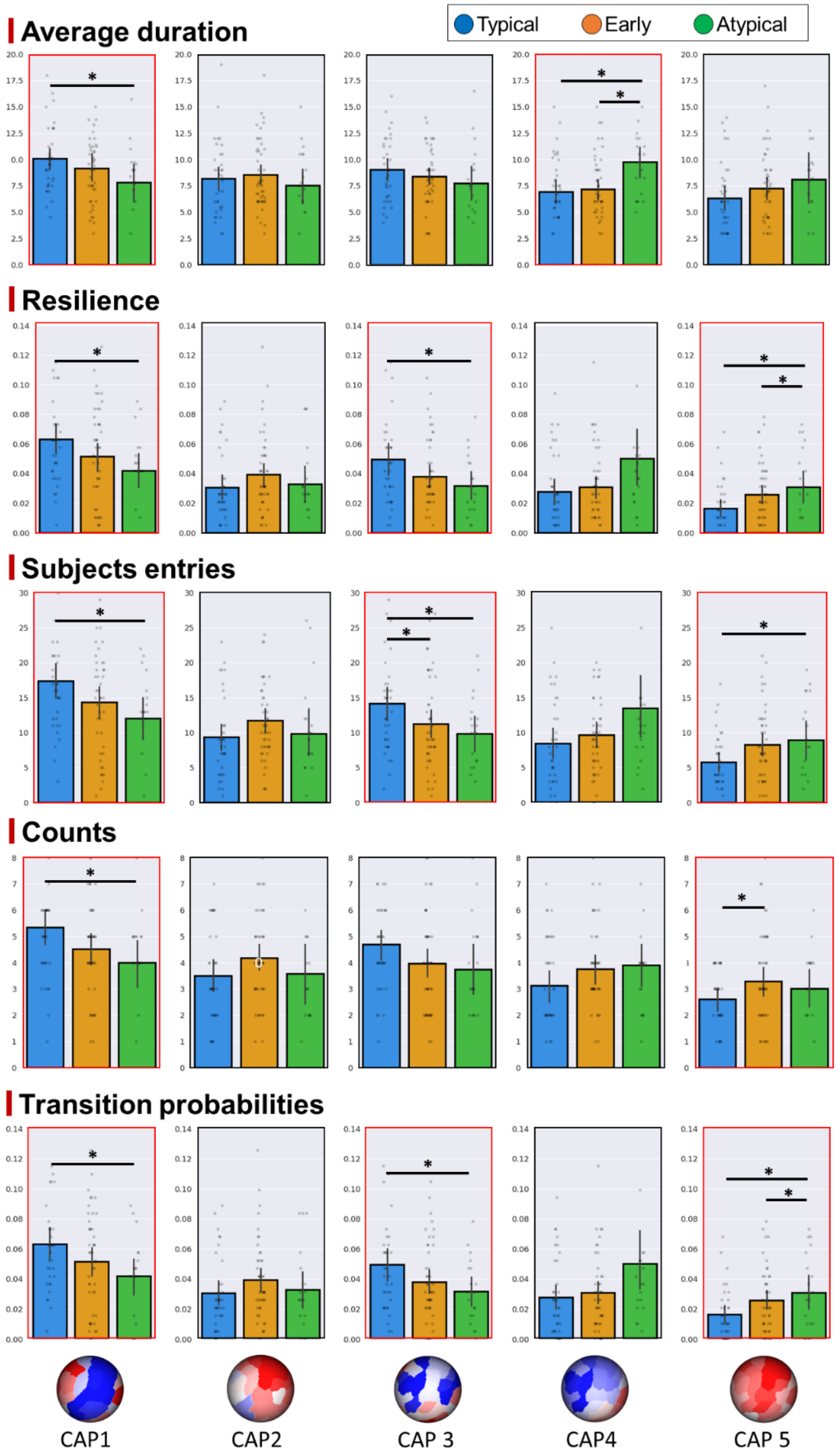


**Figure S5. CAP temporal metrics comparison between patient clusters.**
Data are reported as raw data, while statistical analysis was performed on the age-corrected outcomes. Red boxes highlight significant differences between patients' clusters. * marks post-hoc p<0.05.

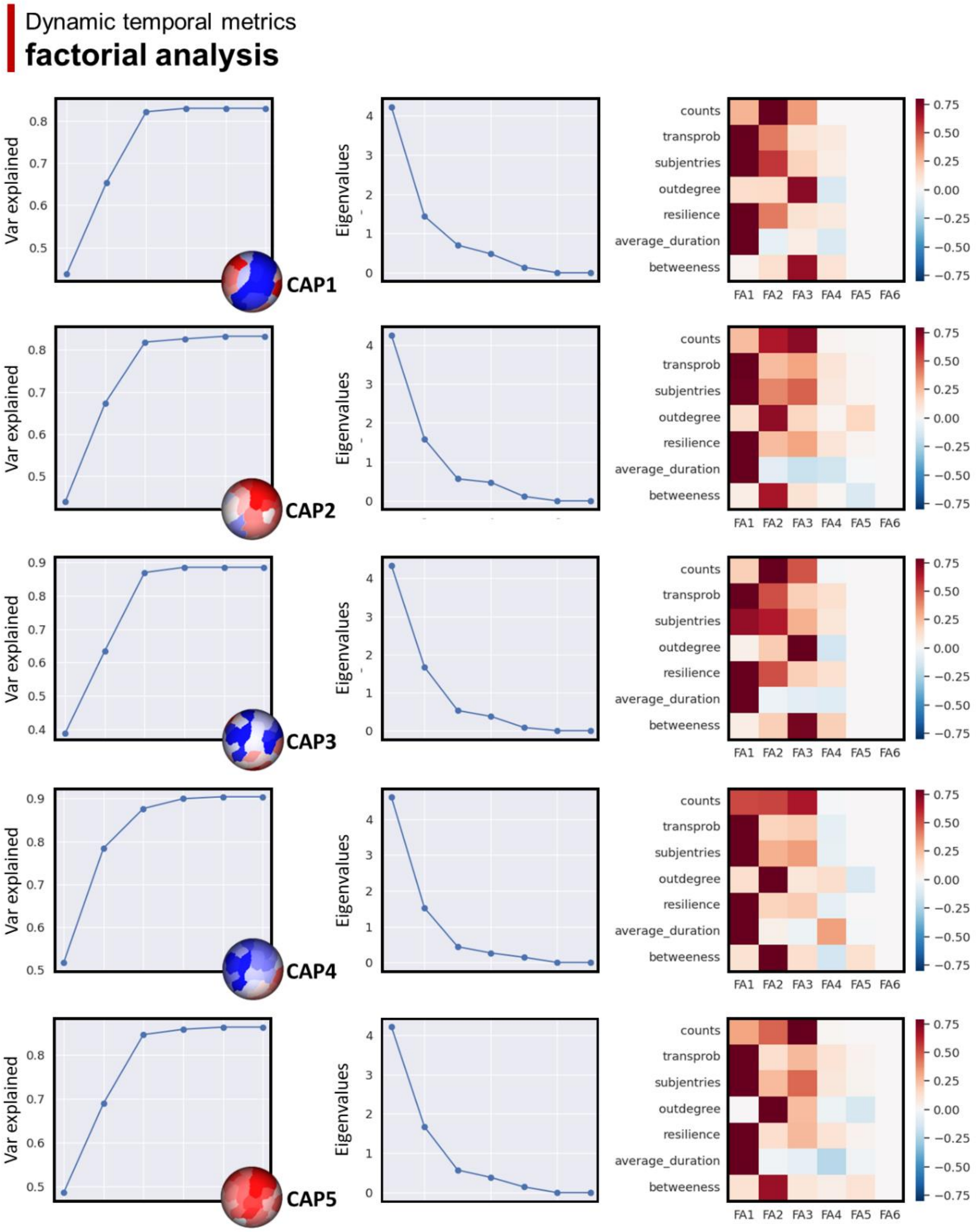


**Figure S6. CAPs temporal dimensionality reduction.**

The factorial analysis identified three main factors for each CAP considered in the analysis. A similar amount of variance (left panels), eigenvalues distribution (middle panels), and loading patterns (right patterns) were reported across CAPs.

### Canonical correlation analysis

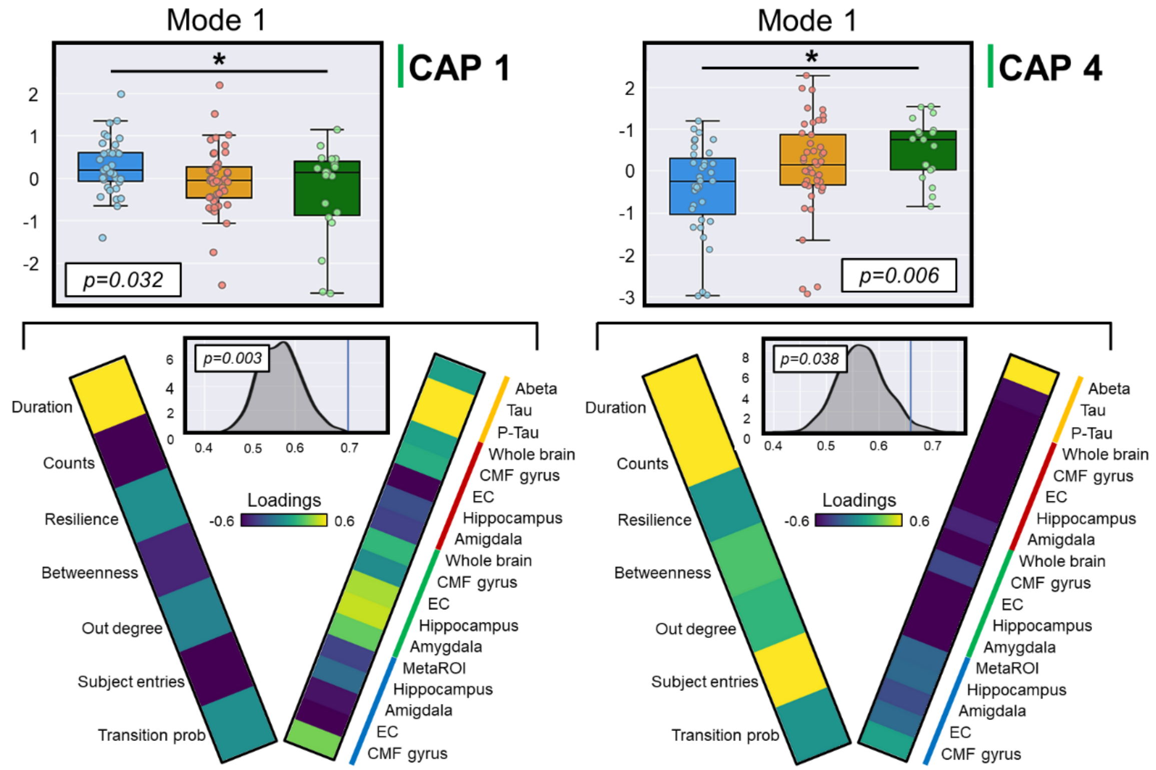


**Figure S7. Results of the canonical correlation analysis.**
CAP1 and CAP4 showed significant modes different between patients' clusters. Post-hoc analysis revealed a significant difference between typical and atypical clusters for both CAP modes. Only modes significant at permutation (gray-distribution plots) and ANOVA (p-values are reported in the white box within each box plot) are shown. Vectors represent the loadings of the first mode for both CAP1 and CAP4 and grouped according to the methodology used to extract the features: CSF (orange line), amyloid PET (red line), Tau PET (green line), and structural MRI (blue line).

**Supplementary Table S1. Sociodemographic, and clinical/cognitive profile of patients' subgroups.** Cognitive composite data (memory, executive functions, language, and visuospatial) are shown as z-score according to the whole ADNI cohort mean and standard deviation distribution (data retrieved directly from the ADNI database).

| | **Typical cluster** | **Early cluster** | **Atypical cluster** | p-value |
|---|---|---|---|---|
| Sex (F) | 49% | 51% | 53% | 0.954 |
| Age (years) | 74.3 ± 6.95 | 70.8 ± 8.33 | 76.4 ± 8.92 | *0.029* |
| Education (years) | 16.6 ± 2.34 | 15.7 ± 2.50 | 16.1 ± 3.33 | 0.351 |
| MMSE | 25.74 ± 2.86 | 28.64 ± 1.55 * | 26.10 ± 2.35 # | *<0.001* |
| CDR-SOB | 3.11 ± 2.32 | 1.07 ± 0.81 * | 2.66 ± 1.89 # | *<0.001* |
| ADAS13 | 23.33 ± 10.40 | 11.43 ± 3.91 * | 18.37 ± 8.74 # | *<0.001* |
| Memory | -0.17 ± 0.71 | 0.74 ± 0.48 * | -0.07 ± 0.72 # | *<0.001* |
| Executive functions | -0.01 ± 1.26 | 0.72 ± 0.81 * | 0.14 ± 0.63 | *0.003* |
| Language | 0.02 ± 1.10 | 0.60 ± 0.68 * | -0.12 ± 0.80 # | *0.002* |
| Visuo-spatial | -0.35 ± 1.03 | -0.01 ± 0.78 | -0.08 ± 0.67 | 0.212 |

### marks differences from the early cluster: * marks significant differences from the typical cluster. Abbreviations: CDR-SOB: clinical dementia rating – sum of boxes; F: female; MMSE: Mini-Mental State Examination.
